# Colorectal cancer screening within colonoscopy capacity constraints: can FIT-based programmes save more lives by trading-off more sensitive test cut-offs against longer screening intervals?

**DOI:** 10.1101/2020.12.17.20242107

**Authors:** Ethna McFerran, James F. O’Mahony, Steffie Naber, Linda Sharp, Ann G. Zauber, Iris Lansdorp-Vogelaar, Frank Kee

## Abstract

**Introduction:** Colorectal cancer (CRC) prevention programmes using faecal immunochemical testing (FIT) as the primary screen typically rely on colonoscopy for secondary and surveillance testing. Colonoscopy capacity is an important constraint, limiting the number of primary tests offered. Many European programmes lack sufficient colonoscopy capacity to provide optimal screening intensity regarding screening age ranges, intervals and FIT cut-offs. It is currently unclear how to optimise programmes within colonoscopy capacity constraints.

**Design:** The MISCAN-Colon microsimulation model was used to determine if more effective CRC screening programmes can be identified within existing colonoscopy capacity. The model assessed 525 strategies of varying screening intervals, age ranges and FIT cut-offs, including previously unevaluated 4 and 5 year screening intervals. These strategies were compared with policy decisions taken in Ireland to provide CRC screening within available colonoscopy capacity. Outcomes estimated net costs, quality-adjusted-life-years and required colonoscopy numbers. The optimal strategies within finite colonoscopy capacity constraints were identified.

**Results:** Combining a reduced FIT cut-off of 10 µg Hb/g, an extended screening interval of 4 years and an age range of 60-72 years requires 6% fewer colonoscopies, reduces net costs by 23% while preventing 15% more CRC deaths and saving 16% more QALYs relative to current policy.

**Conclusion:** Previously overlooked longer screening intervals may balance optimal cancer prevention with finite colonoscopy capacity constraints. Simple changes to screening configurations could save lives, reduce costs, and relieve colonoscopy capacity pressures. These findings are directly relevant to CRC screening programmes across Europe that employ FIT-based testing and face colonoscopy capacity constraints.

## INTRODUCTION

Colorectal cancer (CRC) is a common malignancy that kills approximately 800 000 people globally each year[1]. Early detection improves survival, with survival rates of 90% for locally detected disease versus 13% when metastasized[2]. Screening for CRC has been shown to reduce both incidence and mortality[3]. CRC screening is cost-effective when offered at an appropriate intensity[4,5].

The advent of population-based CRC screening is relatively recent, with 14 EU states only adopting screening after 2009. Organised CRC screening programmes in Europe commonly use faecal-based tests such as the guaiac faecal occult blood tests (gFOBT) or faecal immunochemical testing (FIT)[6]. As of 2015, 20 out of 28 EU member states were in various stages of implementing population-based CRC screening (Table 1)[7]. Recent reports show that more than half of these use FIT[8]. The most common screening interval was every two years, but there are significant differences in FIT thresholds in use, ranging from 6-180 micrograms of haemoglobin per gram of faeces (µg Hb/g).

**Table 1.**
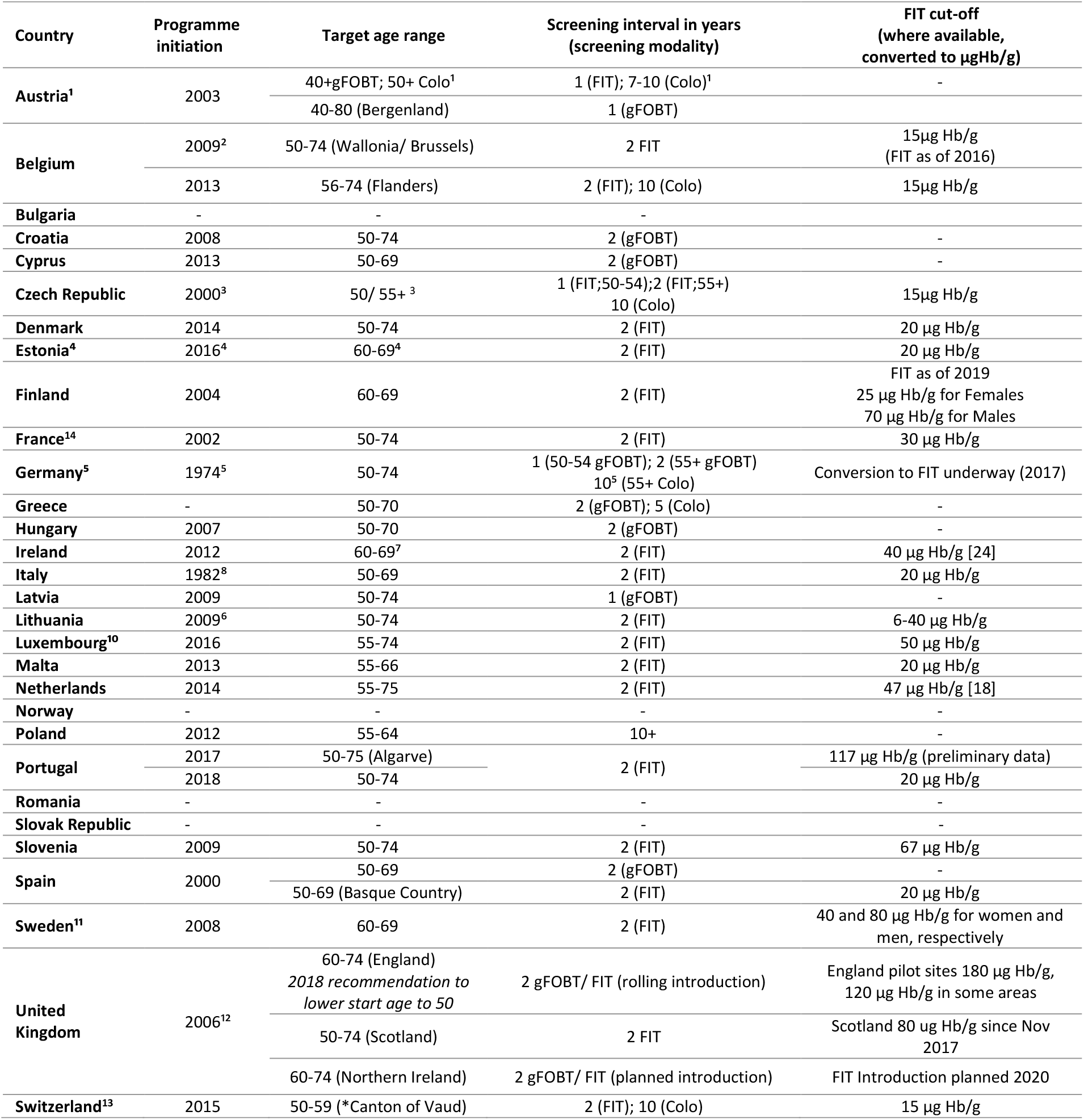
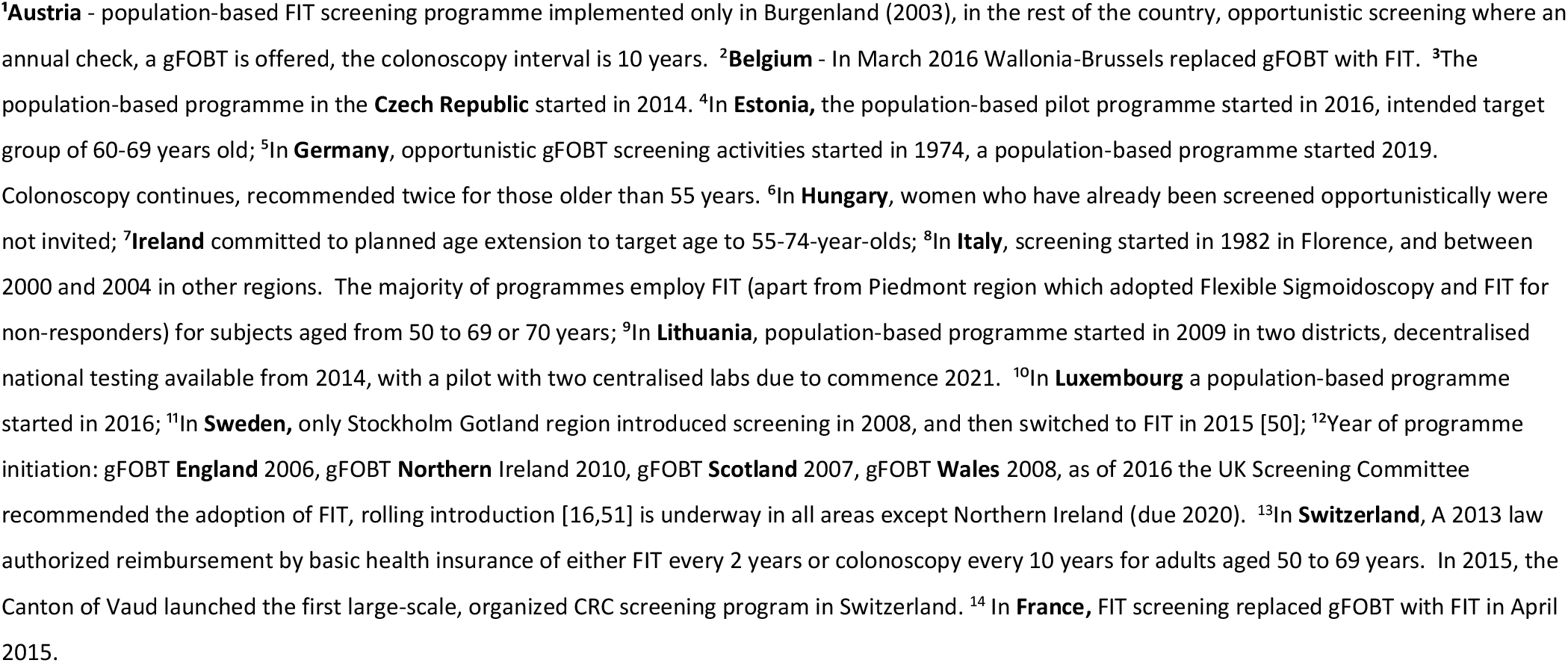
Screening programmes across European countries

Programmes using faecal-based primary screening typically use colonoscopy for secondary diagnostic testing of those with positive screening tests, as well as within alternative routes of referrals and for post-treatment surveillance. Insufficient colonoscopy capacity can constrain what intensity and population coverage of CRC screening is feasible[9,10]. Consequently, colonoscopy capacity imposes limits on how many lives can be saved through CRC screening.

The effectiveness and cost-effectiveness of population CRC screening varies with the breadth of the screening age range and length of the screening interval. In the case of FIT-based testing, effectiveness and cost-effectiveness also vary with the test cut-off used to determine positivity[11]. Reducing the FIT cut-off improves sensitivity at the cost of reduced specificity. Shorter screening intervals, wider screening age ranges and lower FIT cut-offs all lead to increased colonoscopy requirements. Despite the increase in colonoscopies, lower FIT cut-offs are generally more cost-effective[12].

Most cost-effectiveness analyses (CEAs) of CRC screening do not consider the binding colonoscopy capacity constraints. Some studies have, however, shown how finite capacity might be best used in the Dutch context[9,13]. The objective of this study is to further explore the potential for improved effectiveness and cost-effectiveness within a capacity-constrained system. In particular, while most existing CRC screening CEAs have explored screening intervals between 1 and 3 years[14–18], this analysis aims to investigate the potential of longer screening intervals to enhance screening effectiveness within colonoscopy constraints. It uses the example of the policy changes made in the Irish CRC screening programme as a case study to investigate what alternative strategies could improve population health outcomes.

### Case Study

The challenges facing European CRC screening are demonstrated by the case of the Irish CRC screening programme. It serves as a useful example as the screening strategy was initially specified following a health economic analysis and has been modified since in response to colonoscopy capacity constraints. The initial health technology assessment (HTA) that informed the establishment of Ireland’s CRC screening programme was conducted in 2009[19,20]. It simulated comparisons of FIT, gFOBT and once-off sigmoidoscopy. FIT and gFOBT were considered over a limited selection of age ranges at one screening interval of two years. The FIT test performance characteristics were derived from pooled analyses and employed a single FIT cut-off of 20 µg Hb/g of faeces, equivalent to 100 nano-grams of haemoglobin per millilitre of buffer (ng Hb/ml)[20,21]. The HTA found biennial FIT between ages 55-74 was the optimally effective and cost-effective strategy. However, insufficient colonoscopy capacity prevented the implementation of this strategy and prompted further analyses(26, 27). These suggested narrower age range as one way to operate within existing colonoscopy capacity[23]. These subsequent assessments did not examine varying the screening interval or FIT cut-offs.

The programme was launched in October 2012 with biennial screening offered between ages 60-69 at a cut-off of 20 µg Hb/g (FIT 100ng Hb/ml). The stated intention was to expand to the initially planned 55-74 age range as colonoscopy capacity developed[24]. In practice, colonoscopy capacity constraints persisted, leading to a second policy change in early 2014. The FIT cut-off was increased from 20 to 45 µg Hb/g (100 to 225 ng Hb/ml)[25]. While adopting a higher cut-off would improve specificity and ease colonoscopy demand, the loss of sensitivity would reduce screening effectiveness[26]. Restoring the 55-74 age range was recently restated as a policy objective, but reducing the FIT threshold was not[27].

## METHODS

We used a microsimulation model to estimate the costs and effects of a broad range of FIT-based screening strategies. We simulated the policy choices made to date to address colonoscopy capacity constraints and attempted to find alternative policies that are feasible given these constraints but offered greater effectiveness and cost-effectiveness. We used the MISCAN-Colon model to simulate multiple screening strategies in a single birth cohort of average-risk individuals. This established micro-simulation model was developed at the Erasmus University Medical Center[28]. Its underlying structure and parameters have been described elsewhere, alongside comparative evaluations against other CRC screening models[29]. An overview of the model applied in other studies[9,30] is publicly available[28].

MISCAN-Colon simulates the life histories of individuals who may develop one or more adenomas. These adenomas may progress from small (≤5 mm) to medium (6 to 9 mm) to large (≥10 mm) lesions. Some adenomas will develop into preclinical cancer, which may then progress through stages I to IV. Symptomatic presentation of CRC is possible at any stage. Survival after clinical diagnosis is determined by the stage at diagnosis, the localization of the cancer and the person’s age.

Screening alters some of the simulated life histories through the detection and removal of adenomas or the detection of cancer earlier than a clinical presentation, potentially leading to improved prognosis due to earlier treatment[2]. However, screening can also result in serious complications including perforation[31], over-diagnosis and overtreatment of CRC (that is, the detection and treatment of cancer that would not otherwise progress to affect quality of life or life expectancy)[32]. By comparing all simulated life histories with and without screening, MISCAN-Colon estimates the cost and effectiveness of the alternative screening strategies. Whilst patients were not involved in this study due to the nature of the methods applied, this work seeks to advocate for their interests in the policy practice interface.

### Test Characteristics

The FIT test characteristics (Table 2) were taken from published estimates[33]. In the absence of a consistent source of test performance characteristics corresponding to the case study programme cut-off of 45 µg Hb/g (225ng Hb/ml) [20], we used published estimates for 40 µg Hb/g (200 ng Hb/ml) as an approximation. The colonoscopy test characteristics are those applied routinely with MISCAN[34]. The model assumes that 95% of all colonoscopies reach the cecum[35] and that the remaining 5% are distributed uniformly over the colon and rectum.

**Table 2.** Test Characteristics

| FIT Cut-off Level<br>(µg Hb/g)[33]* | Specificity<br>(Per person, %) | Sensitivity per lesion, % |  |  |  |  |
| --- | --- | --- | --- | --- | --- | --- |
|  |  | Adenoma |  |  | CRC |  |
|  |  | ≤5mm | 6-9mm | ≥10mm | CRC early stage | CRC late-stage |
| 10 | 95.79 | 0.0 | 9.6 | 16.1 | 65.0 | 90.0 |
| 15 | 97.05 | 0.0 | 5.7 | 14.4 | 58.5 | 87.0 |
| 20 | 97.76 | 0.0 | 4.4 | 13.1 | 52.0 | 83.5 |
| 30 | 98.34 | 0.0 | 2.9 | 12.3 | 50.5 | 83.0 |
| 40 | 98.70 | 0.0 | 2.5 | 10.3 | 50.0 | 82.5 |
| Colonoscopy [34] |  | 75.0 | 85.0 | 95.0 | 95.0 | 95.0 |

\*according to the manufacturer, the OC-SENSOR delivers 10 mg of faeces into in 2.0 mL of buffer; thus, a test result of 100 ng haemoglobin per mL buffer equals 20 µg haemoglobin per g faeces [21].

### Diagnostic Testing and Surveillance

The model assumes that a diagnostic colonoscopy is offered after any positive FIT. If no adenomas or CRCs are found, individuals return to routine screening at the interval of the simulated strategy. Adenomas detected at colonoscopy are assumed to be removed by polypectomy and individuals then enter colonoscopy-based surveillance following risk-based guidelines: that is, patients received surveillance colonoscopy in 1 and 3-year intervals, in high risk (all lesions ≥10mm) and intermediate-risk (>2 lesions <10mm) respectively[36] to a maximum age of 80 years. Low-risk cases (< 3 adenomas <10mm) are returned to routine FIT screening, based on customary practice[37–39]. The model simulates total colonoscopy requirements for each strategy including those for (secondary) diagnostic testing, surveillance and clinical presentations of the disease.

### Screening, Surveillance Strategies and Attendance Assumptions

As our purpose was to broaden the scope for optimising screening within colonoscopy capacity constraints, we simulated 525 screening strategies in addition to no screening. We modelled five FIT cut-offs of 10, 15, 20, 30 and 40 µg Hb/g (equivalent to 50, 75, 100, 150 and 200 ng Hb/ml). We considered intervals of 1, 2, 3, 4 and 5 years. In addition to the current programme start and stop age of 60 and 70 years respectively, we simulated start ages of 45, 50, 55, 60, 65 and 70 years, with stop ages of 70, 75, 80 years or close approximations thereof depending on the screening interval (Table 3).

**Table 3.** Simulated Screening Strategy Characteristics

| Strategy Characteristics |  |
| --- | --- |
| Screening interval (Years) | 1/2/3/4/5 |
| Start age (Years) | 45/50/55/60/65/70 |
| Stop ages (Years) | 70/75/80 |
| FIT cut-off levels (µg Hb/g) | 10/15/20/30/40 |

All strategies were assessed in terms of the net cost and health effects measured in quality-adjusted life-years (QALYs) relative to no screening. Costs and effects were both discounted at 3% in accordance with the previous Dutch analyses on which our model is based[13,40]. The net costs included the costs of screening, diagnostic colonoscopy, surveillance and any net changes in treatment costs due to early intervention.

We estimated the current colonoscopy capacity constraint in the case study programme as the simulated lifetime colonoscopy demand of the current policy. This was the capacity required for a biennial FIT test in those aged 60-69 with a FIT cut-off of 40 µg Hb/g. We also estimated the implied capacity constraint for the planned expansion of the age range to 55-75 years. We determined the optimally cost-effective strategies within the implied current and future capacity constraints. We used a cost-effectiveness threshold of €20 000/QALY to determine cost-effectiveness[41].

The following results section outlines the overall cost and effect estimates. We give a detailed description of the policy changes taken to date and their estimated outcomes. We then consider what policy alternatives exist within current colonoscopy capacity. Finally, we consider how the programme might be optimally expanded beyond the current colonoscopy capacity.

## RESULTS

An overview of all simulated strategies is presented in Appendix 1, including the FIT cut-off, screening interval and age range along with the estimated colonoscopy requirements, costs, effects and total CRC deaths prevented. Figure 1 plots the estimated costs and effects of all 525 strategies according to FIT cut-off and capacity requirements. The current strategy requires 464 colonoscopies over the lifetime of 1000 individuals. While many strategies exceed current colonoscopy capacity (305 strategies), there are 220 that do not. Many strategies feasible within the colonoscopy constraint (n=85) are more effective than the current strategy (those that lie to the right of the status quo in Figure 1). Some (n=5) also lie below the current strategy, indicating they are cost-saving relative to the current programme. This efficient set is exclusively composed of strategies with a FIT cut-off of 10 µg Hb/g (50 ng Hb/ml). This indicates that the lowest cut-off generally yields strategies that are more effective and less costly.

**Figure 1.**
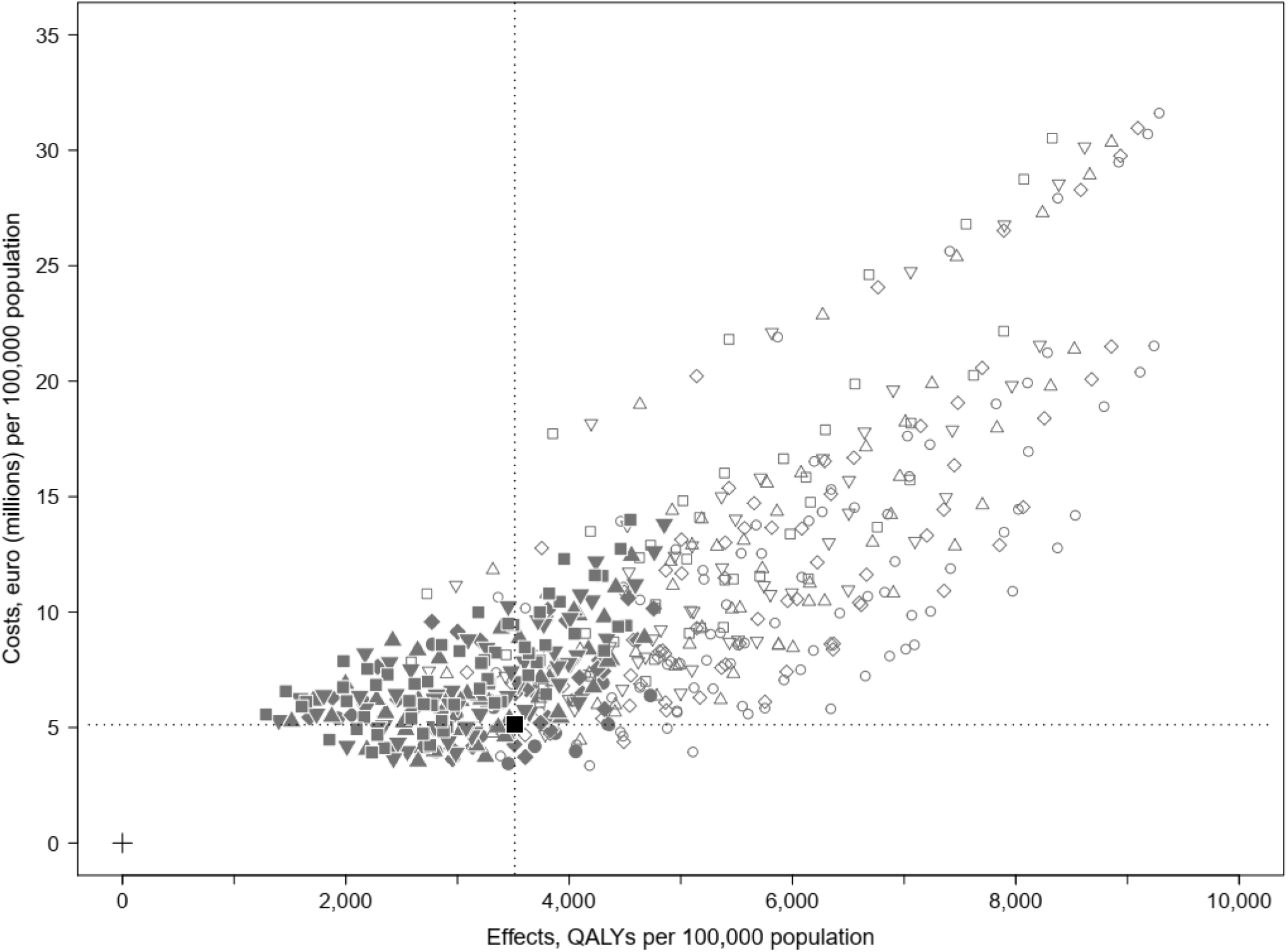
All Strategies by FIT cut-off and colonoscopy capacity

### Decisions reducing colonoscopy capacity requirements

Figure 2 illustrates previous policy changes and some future policy options are detailed in Table 4. The originally HTA-recommended strategy (Point 1) requires a colonoscopy capacity of more than twice the present strategy (1017 colonoscopies per 1000 persons), which would also incur greater costs and yield greater benefits than the status quo. Narrowing the age range to 60-69 adopted at the programme’s introduction in 2012 (Point 2) reduced the colonoscopy requirements by almost half (662 colonoscopies per 1000 persons). The 2014 increase in the FIT cut-off to 45 µg Hb/g further reduces colonoscopy demand, but also reduces effectiveness and modestly increases costs (Point 3: current strategy). The currently planned restoration of the screening age range to 55-75 (Point 4) would generate more QALYs relative to the status quo, however, it would still be less effective than the initially recommended strategy employing the FIT cut-off of 20 µg Hb/g (100 ng Hb/ml) shown by Point 1.

**Table 4.** Summary of Policy Positions

| Identifier | Strategy | Age range | Interval | FIT Cut off<br>(µg Hb/g) | QALYs<br>per 1000 | Cost (€)<br>per 1000 | Colonoscopies<br>per 1000 | Change in<br>QALYs (%)* | Change in<br>Costs (%)* | Change in<br>Colonoscopies<br>(%)* |
| --- | --- | --- | --- | --- | --- | --- | --- | --- | --- | --- |
| 1 | Initial Recommendation | 55-75 | 2 | 20 | 59 | 85,748 | 1,017 | 67 | 67 | 119 |
| 2 | Age restriction | 60-70 | 2 | 20 | 41 | 44,422 | 662 | 17 | -13 | 43 |
| 3 | Approximation of current strategy | 60-70 | 2 | 40 | 35 | 51,201 | 464 | **REF** | **REF** | **REF** |
| 4 | Planned age expansion | 55-75 | 2 | 40 | 52 | 93,152 | 735 | 47 | 82 | 59 |
| A | Max NHB with<br>cost-saving | 60-72 | 4 | 10 | 41 | 39,680 | 437 | 16 | -23 | -6 |
| B | Max NHB within capacity | 55-75 | 5 | 10 | 47 | 63,861 | 455 | 35 | 25 | -2 |
| C | Optimised (Max NHB) with<br>expanded capacity | 50-74 | 4 | 10 | 58 | 95,271 | 707 | 66 | 86 | 52 |
| D | Max overall Net Health Benefit | 50-80 | 1 | 10 | 92 | 215,284 | 3,669 | 163 | 320 | 691 |

\*Percentage change relative to the current strategy (Strategy 3)

**Figure 2.**
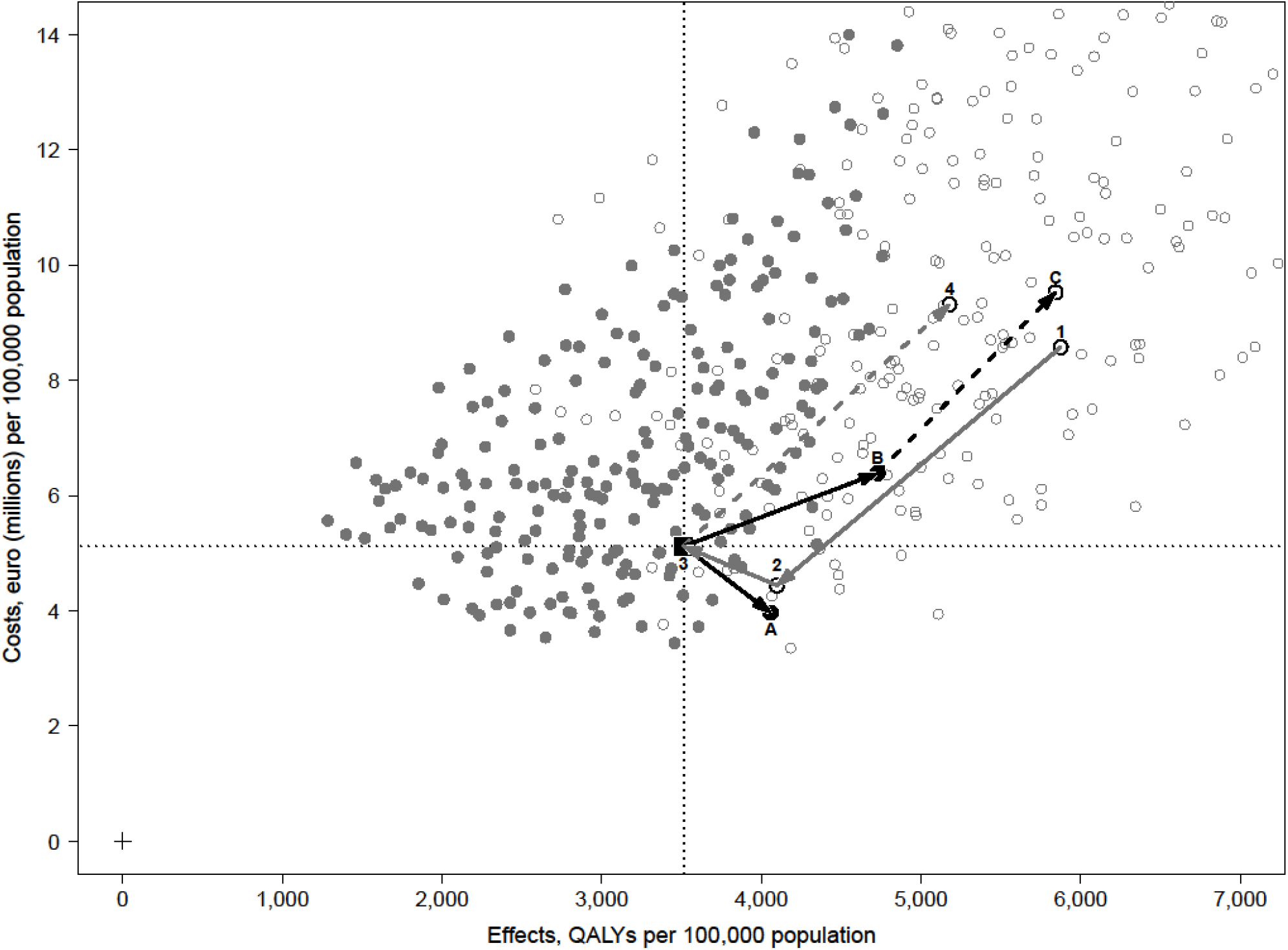
Past Policy Changes and Future Policy Options. Figure 2 shows a subset of Figure 1, with the axes rescaled for clarity. This illustrates previous policy changes chronologically, 1 to 2 and 2 to 3. Future policy options are presented as moves from 3 to A (cost-saving and more effective, requiring 6% fewer colonoscopies.) From 3 to B, providing maximum NHB within capacity. Assuming an increased capacity, as required to provide the planned age expansion point 4, the maximum NHB at that predicted colonoscopy capacity would be provided by point C.

### Potential policy alternatives

Three potential policy alternatives to the status quo are illustrated in Figure 2. Options A and B are both within the current colonoscopy capacity and so are feasible now. Option A uses a FIT cut-off of 10 µg Hb/g (50 ng Hb/ml) with a 4-year screening interval for those aged 60-72. It dominates the current policy, offering 16% more QALYs, 15% more CRC deaths prevented, 23% less costs and requires 6% fewer colonoscopies relative to the current strategy, Strategy 3. Option B is the optimally cost-effective currently feasible strategy. It uses a 10 µg Hb/g (FIT 50 ng Hb/ml) cut-off with a 5-year screening interval between ages 55-75. It provides an approximate 35% gain in QALYs, 29% more CRC deaths prevented, a modest 2% reduction in colonoscopies relative to the current strategy, but at a 25% cost increase.

The current policy commitment to restore the initially-planned 55-74 age range does not mention plans to change the screening interval or FIT cut-off[27]. Consequently, expanding the age range implies a 59% increase in colonoscopy capacity (Point 4). Strategy C is an alternative policy using the same implied increased capacity. This uses a 10 µg Hb/g (FIT 50 ng Hb/ml) cut-off with a 4-year interval between ages 50-74. It would provide a 13% QALY gain relative to the planned age expansion (Strategy 4), but would also be 2% more costly, it would, however, require 4% fewer colonoscopies than those predicted for the planned age expansion.

Finally, Figure 3 shows the overall optimally cost-effective strategy without any colonoscopy capacity constraint Point D which uses annual screening between ages 50-80 at a FIT cut-off 10 µg Hb/g (50 ng Hb/ml). This would require a considerable increase in colonoscopy capacity of 691% relative to the status quo and would cost 320% more but would yield an estimated 163% more QALYs and 111% more CRC deaths prevented.

**Figure 3.**
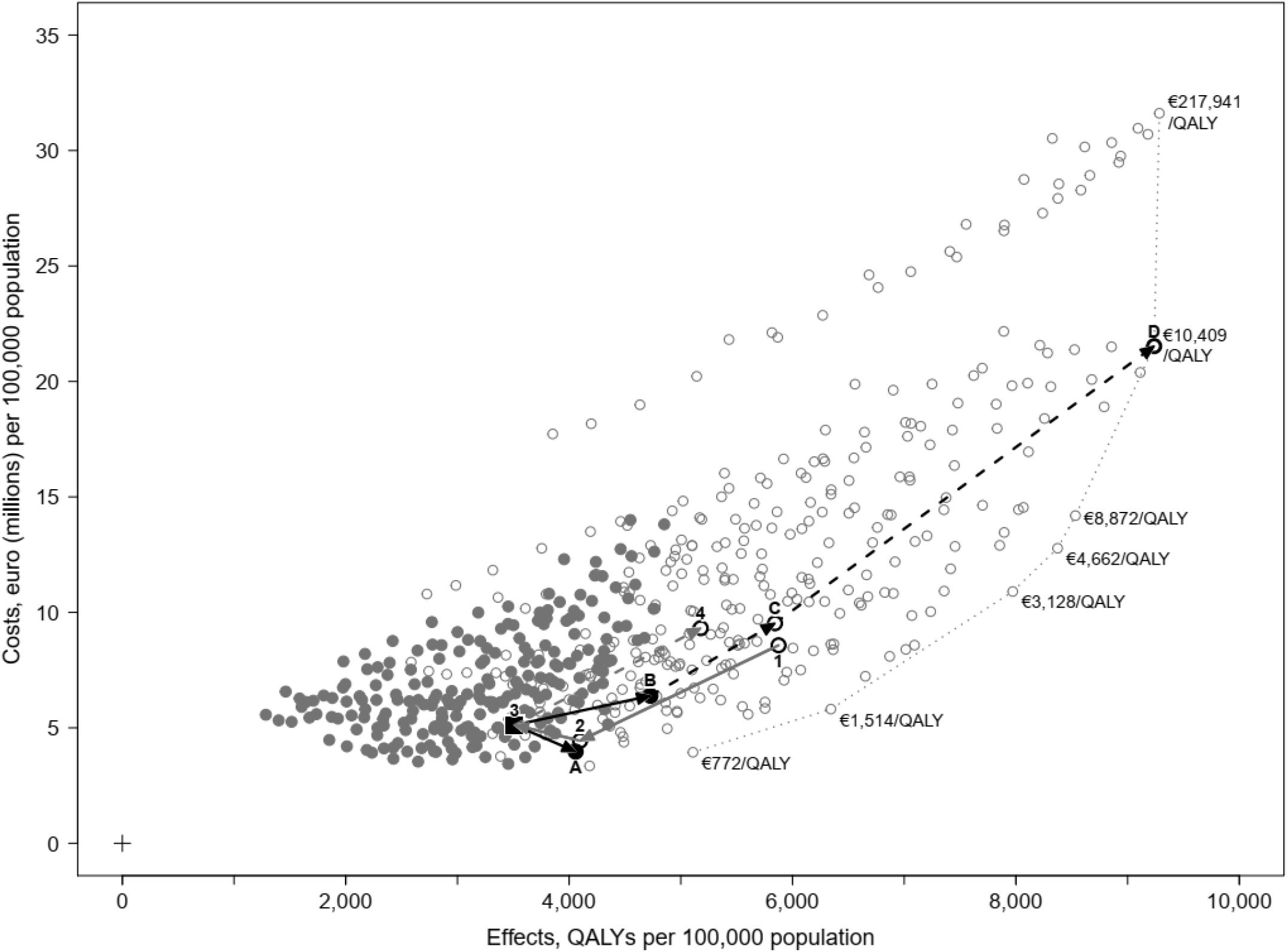
Policy Overview, including overall Max NHB policy (D)

## DISCUSSION

Our analysis shows that the optimal policy response to limited colonoscopy capacity may not be to raise the FIT cut-off level, or widen the screening age-range, but rather to use longer screening intervals and more sensitive cut-off levels. In our case study, the policy response to limited capacity to date has been to preserve biennial screening while narrowing the screening age range and raising the FIT cut-off. Modelling indicates that this runs counter to what makes the most effective use of scarce colonoscopy services. We find that by lengthening the screening interval we can maintain a broad screening age range and retain a more sensitive FIT cut-off and deliver greater benefits in terms of CRC deaths prevented. Costs would also be reduced by this approach. The primary explanation for our findings is the diminishing marginal returns of intensifying the frequency of screening: screening more people less often with a more sensitive FIT threshold seems a more efficient way of reducing the colonoscopy requirements than screening fewer people more frequently with a less sensitive FIT threshold.

Our findings are of clear policy relevance to the many countries facing difficulties in implementing CRC screening within constrained colonoscopy capacity, especially following the initial introduction of national programmes[42]. Restricting the screening age range and reducing the positivity threshold sensitivity of FIT appears a common policy response. A recent EU review of cancer screening services noted that “to optimise (limited) resource allocation, by maximising the cost-effectiveness ratio of the intervention, and to match their endoscopy capacity, several EU member states had actually adopted screening policies targeting a stricter age range, usually shifted to the older age groups, showing a higher prevalence of disease, resulting in a lower cost per lesion detected”[7]. Our results raise the possibility that these countries may also be making what seem like logical, but potentially sub-optimal policy responses to capacity constraints.

Our results differ from other studies of CRC screening using the same MISCAN-Colon model to estimate the optimal policy response to scarce colonoscopy capacity[9,13]. They found higher FIT cut-offs would be optimal under binding colonoscopy constraints. Our results differ from Van Hees et al. as, among other reasons, that analysis addressed a set of alternatives within an already limited screening age range as set out by policymakers [9]. Similarly, our conclusions differ from Wilschut et al. because we simulated a broader range of screening intervals including every 4 and 5 years[13]. As such, our analysis adds a novel finding to the literature on optimal CRC screening within constrained colonoscopy capacity.

Our findings illustrate the general principle that a cancer screening CEA should simulate a broad range of policy alternatives to find the optimal strategy. The initial HTA within the case study assessed only a small range of strategies and was published prior to work showing the benefit of varying FIT cut-offs. This led that analysis to overlook the issue of diminishing marginal returns of shortening the screening interval. Accordingly, it could not identify the benefits of applying longer intervals to more people, rather than retaining short intervals for a narrow age range. Whilst the original HTA was supplemented by additional evaluations, these too did not consider strategies with longer intervals[24].

Within the specific context of Ireland, better CRC prevention will require careful planning. A similar conclusion could, therefore, apply to many other European countries. The Irish Cancer Society has raised concerns regarding colonoscopy capacity constraints and the emergence of inequities of access to colonoscopy between public and private patients[43,44]. The recently renewed National Cancer Strategy restated the plan to expand capacity to permit the restoration of the screening age range to 55-74 by end-2021[27]. Plans for this ambitious capacity expansion are being managed by Ireland’s Health Service Executive National Endoscopy Steering Group[27].

While the currently planned expansion of colonoscopy capacity is welcome, our results indicate that the case study programme will remain unnecessarily inefficient. Modelling suggests that considerable improvements could be achieved if longer intervals of 4 years were adopted instead of the current 2-year interval. An increase in the screening interval could lead decision-makers to worry that the public might become confused and adherence could suffer. While such potential concerns are understandable, there is no evidence that adherence would be compromised, given the use of wider intervals in other disease areas. Conversely, the modelling evidence suggests that persisting with the present policy is likely to save fewer lives than other feasible strategies.

Our results also highlight a broader concern about the sufficiency of CRC screening programmes in Ireland and other European nations. While Ireland plans to expand colonoscopy capacity, the current policy commitment still falls far short of what is ultimately required. Our results indicate that much larger gains could be made if annual screening were adopted, while remaining cost-effective (Strategy D, Figure 3). This again emphasises the need for CEAs to consider a broad range of options. Current recommendations for biennial screening throughout Europe might lead policymakers to accept very considerable under-provision of CRC screening and save too few lives.

Our analysis naturally has some limitations. Firstly, to date, no trial or observational data has examined the long-term effect of varying FIT intervals[45], thus the correlation between multiple tests and absolute risk, especially in the context of non-bleeding lesions, remains uncertain. Accordingly, the conclusions presented here on both extending the interval and using annual screening depend heavily on the current model assumptions. More data might be required to give decision-makers confidence in varying the screening interval. Despite this, our analysis usefully illustrates what additional studies could be beneficial to undertake. Secondly, the model reflects Dutch disease incidence and healthcare costs and therefore can only give a broad indication of what is likely to apply in an Irish context. Furthermore, in common with many screening HTAs, we assumed 100% screening adherence. Currently, uptake within the national bowel cancer screening programme is approximately 40%[24]. Similarly, the FIT cut-off of 45 µg Hb/g as used in the programme would generate fewer false positives than we inferred by using a 40 µg Hb/g cut-off. Consequently, our analysis may marginally over-estimate current colonoscopy capacity. However, this approximation was necessary given the need for a consistent source for the test performance characteristics of the alternative FIT cut-offs. Finally, the model assumes 95% of colonoscopies reach the caecum. This may overestimate the effectiveness of the procedure as studies have shown that this can be lower[46,47]. Despite these simplifications, we are confident that the analysis valuably illustrates the relevance of considering a broad range of policy alternatives and a clear indication of how a national bowel cancer screening programme could save more lives.

An explicit acknowledgement of the relevance of the COVID-19 pandemic to our study is necessary. Our analysis was conceived before the advent of COVID-19 and does not reflect the additional capacity challenges that population-based CRC screening programmes are now facing as they attempt to mitigate transmission risk. Our study naturally does not reflect these additional constraints, but the principles of our findings remain the same. Indeed, the possibility that capacity constraints in CRC screening will be exacerbated in the medium term heighten the relevance of our conclusions.

Adopting annual FIT would require very large increases in colonoscopy capacity for many countries. In the Irish context, we suggest that a revision of the HTA evidence supporting the CRC screening programme is now timely, both for the medium-term optimisation of current capacity and the longer-term planning of overall colonoscopy capacity requirements. It is now necessary to revisit and expand previous analyses of CRC screening and consider additional policy alternatives. Such evidence and policy reviews are now required elsewhere in Europe too. Given that trials examining the effectiveness of FIT may not be available for another ten years[48] modelling provides for more timely improvements. Given the interim shortfall in trial data, initiatives such as the EUTOPIA screening modelling project will be useful in assisting member states to inform such reviews[49].

## CONCLUSION

Existing CRC screening programmes may be unnecessarily ineffective and inefficient if analyses informing their design do not consider a wide range of strategies. In our case study, more lives and health services costs could be saved within existing colonoscopy capacity constraints if a lengthening of the screening interval was traded-off against an increase in population coverage and the adoption of a more sensitive FIT cut-off. A broader finding is that much larger increases in colorectal cancer screening capacity than is currently planned appears warranted if annual screening were to be adopted. Policymakers must recognise the need to consider all policy alternatives, within both current colonoscopy capacity constraints and future expanded service capacity. Otherwise, many avoidable colorectal cancer deaths will result over the coming decades. The findings from this case study are likely to be highly relevant for all European nations implementing FIT-based CRC screening with biennial intervals in the face of constrained colonoscopy capacity.

## Data Availability

All data generated by this research has been provided, for further enquiries please contact the corresponding author.

## ADDITIONAL INFORMATION

The authors declare no conflict of interest.

## Authors’ contributions

EMF and JFOM conducted the simulation modelling experiments and wrote the manuscript. SN, LS, FK and AGZ helped review and edit the manuscript. ILV supported MISCAN model access, provided technical advice on study design, and manuscript review.

## Acknowledgements

Thanks to Irene Waters at the Irish National Screening Service for providing input on the Bowel Screen programme.

## FIGURES LEGENDS

Strategies with FIT cut-offs of 10, 15, 20, 30 and 40 µg Hb/g are shown with round, triangular, diamond, inverted triangular and square markers respectively. Strategies that are within the colonoscopy capacity of the current strategy are shown with solid markers, whereas those exceeding current capacity are shown with hollow markers. The current strategy is shown as the black square. The black dotted lines correspond to the costs and effects of the status quo. The efficient frontier is shown with the grey dotted line.

Figure 1 Legend

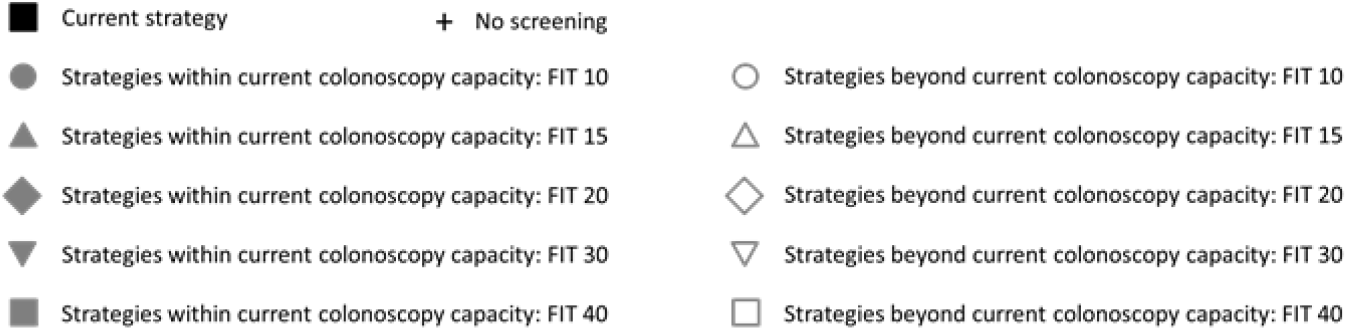

Figure 2 Legend

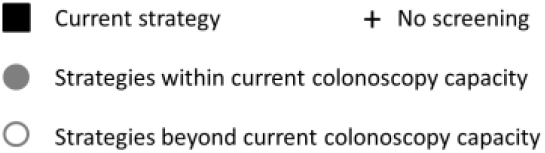

Figure 3 Legend

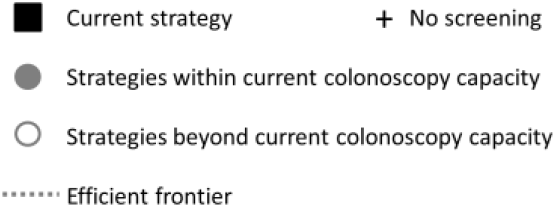

## Appendices

**APPENDIX 1.** Results of All Strategies Modelled (sorted by FIT cut-off)

| Strategy Characteristics |  |  |  |  |  | Estimated Outcomes (for 10 million persons simulated) |  |  |  | Comparisons Relative to Current Policy |  |  |  | Total CRC deaths prevented | ICER |
| --- | --- | --- | --- | --- | --- | --- | --- | --- | --- | --- | --- | --- | --- | --- | --- |
| Policy Position | Cut off (µg Hb/g) | Interval (Years) | Start Age | Stop Age | Number of Lifetime Screens | ΔQALY | ΔC (€) | ΔLYG | Total Number of Colonoscopies | Within Capacity 0= No 1= Yes | % ΔQALY | % ΔC | %ΔNo. Colonoscopies |  |  |
|  | 0 | 0 | 0 | 0 | 0 | 0 | 0 | 0 | 0 |  |  |  |  | 0 | - |
| Max overall NHB (D) | 10 | 1 | 50 | 80 | 30 | 923,774 | 2,152,841,980 | 962,802 | 36,690,721 | 0 | 163 | 320 | 691 | 91186 | 10409 |
|  | 10 | 1 | 50 | 75 | 25 | 911,291 | 2,038,845,024 | 948,937 | 36,029,947 | 0 | 159 | 298 | 677 | 88557 | - |
|  | 10 | 1 | 50 | 70 | 20 | 878,990 | 1,889,923,187 | 914,974 | 34,438,993 | 0 | 150 | 269 | 643 | 83408 | - |
|  | 10 | 1 | 55 | 80 | 25 | 853,279 | 1,419,041,116 | 892,270 | 32,412,179 | 0 | 143 | 177 | 599 | 90489 | 8872 |
|  | 10 | 1 | 55 | 75 | 20 | 837,341 | 1,277,636,957 | 874,613 | 31,587,240 | 0 | 138 | 150 | 581 | 87173 | 4662 |
|  | 10 | 1 | 45 | 80 | 35 | 928,401 | 3,161,158,158 | 969,447 | 38,998,864 | 0 | 164 | 517 | 741 | 87349 | 217941 |
|  | 10 | 1 | 45 | 75 | 30 | 918,185 | 3,070,154,354 | 958,158 | 38,465,302 | 0 | 161 | 500 | 729 | 85224 | - |
|  | 10 | 1 | 45 | 70 | 25 | 892,149 | 2,948,308,364 | 930,661 | 37,189,552 | 0 | 154 | 476 | 702 | 81081 | - |
|  | 10 | 1 | 55 | 70 | 15 | 797,157 | 1,090,283,748 | 832,305 | 29,563,393 | 0 | 127 | 113 | 538 | 80744 | 3128 |
|  | 10 | 1 | 50 | 65 | 15 | 811,243 | 1,695,206,231 | 845,463 | 31,394,594 | 0 | 131 | 231 | 577 | 74679 | - |
|  | 10 | 1 | 45 | 65 | 20 | 837,582 | 2,792,411,142 | 874,754 | 34,789,084 | 0 | 138 | 445 | 650 | 74040 | - |
|  | 10 | 1 | 55 | 65 | 10 | 709,224 | 857,943,179 | 742,492 | 25,578,155 | 0 | 102 | 68 | 452 | 69389 | - |
|  | 10 | 1 | 60 | 80 | 20 | 706,852 | 986,260,798 | 749,691 | 26,540,438 | 0 | 101 | 93 | 472 | 83750 | - |
|  | 10 | 1 | 60 | 75 | 15 | 686,998 | 809,446,643 | 727,648 | 25,490,509 | 0 | 96 | 58 | 450 | 79597 | - |
|  | 10 | 1 | 50 | 60 | 10 | 685,214 | 1,423,396,387 | 718,065 | 26,200,406 | 0 | 95 | 178 | 465 | 61184 | - |
|  | 10 | 1 | 45 | 60 | 15 | 740,870 | 2,563,015,440 | 776,495 | 30,771,718 | 0 | 111 | 401 | 564 | 63639 | - |
|  | 10 | 1 | 60 | 70 | 10 | 634,347 | 581,066,953 | 672,429 | 22,818,047 | 0 | 81 | 13 | 392 | 71121 | 1514 |
|  | 10 | 1 | 65 | 80 | 15 | 523,101 | 791,176,426 | 570,808 | 19,654,061 | 0 | 49 | 55 | 324 | 72051 | - |
|  | 10 | 1 | 45 | 55 | 10 | 586,770 | 2,190,946,536 | 620,525 | 24,617,684 | 0 | 67 | 328 | 431 | 49193 | - |
|  | 10 | 1 | 65 | 75 | 10 | 496,510 | 571,947,109 | 541,520 | 18,263,474 | 0 | 41 | 12 | 294 | 66537 | - |
|  | 10 | 1 | 70 | 80 | 10 | 334,539 | 738,080,543 | 385,218 | 12,362,713 | 0 | -5 | 44 | 167 | 56490 | - |
|  | 10 | 2 | 50 | 80 | 15 | 802,314 | 1,444,752,482 | 872,082 | 17,539,998 | 0 | 128 | 182 | 278 | 84887 | - |
|  | 10 | 2 | 45 | 81 | 18 | 828,387 | 2,123,170,365 | 900,496 | 19,368,532 | 0 | 136 | 315 | 318 | 84566 | - |
|  | 10 | 2 | 50 | 76 | 13 | 789,494 | 1,345,952,060 | 855,800 | 17,124,596 | 0 | 125 | 163 | 269 | 81704 | - |
|  | 10 | 2 | 45 | 75 | 15 | 810,525 | 1,992,527,265 | 878,106 | 18,804,614 | 0 | 131 | 289 | 306 | 80251 | - |
|  | 10 | 2 | 45 | 71 | 13 | 782,377 | 1,901,606,608 | 845,891 | 18,089,982 | 0 | 123 | 271 | 290 | 75224 | - |
|  | 10 | 2 | 50 | 70 | 10 | 741,534 | 1,188,515,223 | 800,689 | 15,895,718 | 0 | 111 | 132 | 243 | 73277 | - |
|  | 10 | 2 | 55 | 81 | 13 | 723,695 | 1,003,088,247 | 792,025 | 15,090,209 | 0 | 106 | 96 | 225 | 82307 | - |
| Policy Position | Cut off (µg Hb/g) | Interval (Years) | Start Age | Stop Age | Number of Lifetime Screens | ΔQALY | ΔC (€) | ΔLYG | Total Number of Colonoscopies | Within Capacity 0= No 1= Yes | % ΔQALY | % ΔC | %ΔNo. Colonoscopies |  |  |
|  | 10 | 2 | 55 | 75 | 10 | 701,455 | 839,544,597 | 764,056 | 14,372,371 | 0 | 100 | 64 | 210 | 76944 | - |
|  | 10 | 2 | 55 | 71 | 8 | 665,232 | 723,266,647 | 722,585 | 13,441,645 | 0 | 89 | 41 | 190 | 70506 | - |
|  | 10 | 2 | 50 | 66 | 8 | 682,599 | 1,086,265,340 | 736,108 | 14,526,217 | 0 | 94 | 112 | 213 | 65015 | - |
|  | 10 | 2 | 45 | 65 | 10 | 702,995 | 1,762,301,445 | 759,134 | 16,296,739 | 0 | 100 | 244 | 251 | 64317 | - |
|  | 10 | 2 | 60 | 80 | 10 | 592,420 | 705,902,275 | 658,520 | 12,035,801 | 0 | 69 | 38 | 160 | 74578 | - |
|  | 10 | 2 | 60 | 76 | 8 | 575,371 | 583,723,782 | 637,209 | 11,493,660 | 0 | 64 | 14 | 148 | 70415 | - |
|  | 10 | 2 | 45 | 61 | 8 | 619,463 | 1,652,385,064 | 670,127 | 14,509,698 | 0 | 76 | 223 | 213 | 54906 | - |
|  | 10 | 2 | 55 | 65 | 5 | 560,257 | 558,610,770 | 608,155 | 11,004,228 | 0 | 59 | 9 | 137 | 55976 | - |
|  | 10 | 2 | 60 | 70 | 5 | 510,850 | 394,125,332 | 563,686 | 9,804,082 | 0 | 45 | -23 | 111 | 59017 | 772 |
|  | 10 | 2 | 50 | 60 | 5 | 535,459 | 909,694,154 | 579,400 | 11,372,549 | 0 | 52 | 78 | 145 | 48455 | - |
|  | 10 | 2 | 65 | 81 | 8 | 438,231 | 629,411,196 | 502,881 | 8,984,357 | 0 | 25 | 23 | 94 | 64417 | - |
|  | 10 | 2 | 65 | 75 | 5 | 406,673 | 425,304,283 | 464,060 | 7,979,930 | 0 | 16 | -17 | 72 | 56887 | - |
|  | 10 | 2 | 45 | 55 | 5 | 446,112 | 1,393,734,864 | 486,162 | 10,823,546 | 0 | 27 | 172 | 133 | 37968 | - |
|  | 10 | 2 | 70 | 80 | 5 | 275,209 | 603,227,178 | 334,639 | 5,650,250 | 0* | -22 | 18 | 22 | 49100 | - |
|  | 10 | 3 | 45 | 81 | 12 | 723,132 | 1,725,591,172 | 807,480 | 11,959,541 | 0 | 106 | 237 | 158 | 77229 | - |
|  | 10 | 3 | 50 | 80 | 10 | 691,806 | 1,219,123,010 | 774,726 | 10,834,242 | 0 | 97 | 138 | 134 | 76616 | - |
|  | 10 | 3 | 45 | 75 | 10 | 704,712 | 1,586,774,580 | 782,403 | 11,479,280 | 0 | 101 | 210 | 148 | 72326 | - |
|  | 10 | 3 | 50 | 74 | 8 | 667,403 | 1,068,268,766 | 742,491 | 10,245,532 | 0 | 90 | 109 | 121 | 70565 | - |
|  | 10 | 3 | 50 | 71 | 7 | 642,359 | 995,380,953 | 712,249 | 9,780,945 | 0 | 83 | 94 | 111 | 65894 | - |
|  | 10 | 3 | 45 | 69 | 8 | 655,535 | 1,452,838,258 | 723,613 | 10,624,442 | 0 | 87 | 184 | 129 | 63539 | - |
|  | 10 | 3 | 55 | 79 | 8 | 618,771 | 833,750,463 | 696,773 | 9,201,050 | 0 | 76 | 63 | 98 | 72665 | - |
|  | 10 | 3 | 55 | 76 | 7 | 607,300 | 750,310,851 | 681,183 | 8,901,165 | 0 | 73 | 47 | 92 | 69630 | - |
|  | 10 | 3 | 45 | 66 | 7 | 614,689 | 1,394,869,637 | 677,239 | 10,004,383 | 0 | 75 | 172 | 116 | 57774 | - |
|  | 10 | 3 | 55 | 70 | 5 | 555,147 | 592,314,553 | 618,020 | 7,901,249 | 0 | 58 | 16 | 70 | 59730 | - |
|  | 10 | 3 | 50 | 65 | 5 | 557,050 | 865,773,932 | 614,793 | 8,406,382 | 0 | 59 | 69 | 81 | 53501 | - |
|  | 10 | 3 | 60 | 81 | 7 | 511,883 | 672,359,371 | 587,074 | 7,349,506 | 0* | 46 | 31 | 58 | 67378 | - |
|  | 10 | 3 | 60 | 75 | 5 | 487,855 | 496,380,554 | 554,500 | 6,712,099 | 0* | 39 | -3 | 45 | 60979 | - |
|  | 10 | 3 | 45 | 60 | 5 | 495,488 | 1,271,389,179 | 546,082 | 8,306,459 | 0 | 41 | 148 | 79 | 43868 | - |
|  | 10 | 3 | 55 | 64 | 3 | 448,274 | 462,275,645 | 497,758 | 6,212,031 | 0* | 28 | -10 | 34 | 44965 | - |
|  | 10 | 3 | 60 | 69 | 3 | 418,359 | 335,033,755 | 472,381 | 5,500,475 | 0* | 19 | -35 | 19 | 48630 | - |
|  | 10 | 3 | 50 | 59 | 3 | 414,833 | 728,871,279 | 459,007 | 6,349,948 | 0* | 18 | 42 | 37 | 37485 | - |
| Policy Position | Cut off (µg Hb/g) | Interval (Years) | Start Age | Stop Age | Number of Lifetime Screens | ΔQALY | ΔC (€) | ΔLYG | Total Number of Colonoscopies | Within Capacity 0= No 1= Yes | % ΔQALY | % ΔC | %ΔNo. Colonoscopies |  |  |
|  | 10 | 3 | 65 | 80 | 5 | 373,947 | 569,414,230 | 443,045 | 5,496,182 | 0* | 6 | 11 | 19 | 56626 | - |
|  | 10 | 3 | 65 | 74 | 3 | 338,594 | 376,439,372 | 397,295 | 4,657,950 | 0* | -4 | -26 | 0 | 48047 | - |
|  | 10 | 3 | 45 | 54 | 3 | 336,341 | 1,064,714,167 | 373,269 | 5,983,644 | 0* | -4 | 108 | 29 | 28372 | - |
|  | 10 | 3 | 70 | 79 | 3 | 233,446 | 537,708,524 | 293,457 | 3,647,597 | 1* | -34 | 5 | -21 | 42708 | - |
|  | 10 | 4 | 45 | 81 | 9 | 634,650 | 1,531,006,698 | 723,683 | 8,436,251 | 0 | 81 | 199 | 82 | 70096 | - |
|  | 10 | 4 | 45 | 77 | 8 | 626,646 | 1,434,333,688 | 710,842 | 8,176,163 | 0 | 78 | 180 | 76 | 67303 | - |
|  | 10 | 4 | 50 | 82 | 8 | 608,340 | 1,151,702,200 | 696,295 | 7,668,753 | 0 | 73 | 125 | 65 | 70213 | - |
| Optimised (Max NHB) with expanded capacity (C) | 10 | 4 | 50 | 74 | 6 | 584,285 | 952,705,745 | 661,351 | 7,065,385 | 0* | 66 | 86 | 52 | 63306 | - |
|  | 10 | 4 | 45 | 69 | 6 | 572,330 | 1,253,621,886 | 642,319 | 7,358,045 | 0 | 63 | 145 | 59 | 56515 | - |
|  | 10 | 4 | 50 | 70 | 5 | 551,155 | 857,413,543 | 619,985 | 6,583,323 | 0* | 57 | 67 | 42 | 56997 | - |
|  | 10 | 4 | 55 | 79 | 6 | 544,582 | 776,942,952 | 625,176 | 6,502,716 | 0* | 55 | 52 | 40 | 65848 | - |
|  | 10 | 4 | 55 | 75 | 5 | 528,949 | 668,137,370 | 603,165 | 6,145,744 | 0* | 51 | 30 | 33 | 61562 | - |
|  | 10 | 4 | 55 | 71 | 4 | 496,870 | 565,688,439 | 562,802 | 5,638,202 | 0* | 41 | 10 | 22 | 55137 | - |
|  | 10 | 4 | 45 | 65 | 5 | 519,871 | 1,181,088,410 | 581,174 | 6,735,302 | 0* | 48 | 131 | 45 | 48901 | - |
|  | 10 | 4 | 50 | 66 | 4 | 499,193 | 777,776,913 | 558,903 | 5,939,650 | 0* | 42 | 52 | 28 | 49088 | - |
|  | 10 | 4 | 60 | 80 | 5 | 463,505 | 673,509,708 | 540,643 | 5,576,630 | 0* | 32 | 32 | 20 | 63020 | - |
|  | 10 | 4 | 55 | 67 | 3 | 446,037 | 480,511,662 | 502,330 | 4,955,736 | 0* | 27 | -6 | 7 | 46886 | - |
|  | 10 | 4 | 60 | 76 | 4 | 436,522 | 507,372,483 | 506,403 | 4,870,522 | 0* | 24 | -1 | 5 | 56323 | - |
|  | 10 | 4 | 45 | 61 | 4 | 448,757 | 1,108,387,476 | 500,926 | 5,947,920 | 0* | 28 | 116 | 28 | 40349 | - |
|  | 10 | 4 | 50 | 62 | 3 | 426,384 | 706,498,688 | 476,562 | 5,116,824 | 0* | 21 | 38 | 10 | 39950 | - |
| Max NHB with cost saving (A) | 10 | 4 | 60 | 72 | 3 | 405,847 | 396,797,580 | 467,287 | 4,368,403 | 1* | 16 | -23 | -6 | 49801 | - |
|  | 10 | 4 | 45 | 57 | 3 | 360,844 | 1,016,977,789 | 403,308 | 4,981,988 | 0* | 3 | 99 | 7 | 31139 | - |
|  | 10 | 4 | 65 | 81 | 4 | 332,405 | 587,973,994 | 403,421 | 3,962,115 | 1* | -5 | 15 | -15 | 52048 | - |
|  | 10 | 4 | 65 | 77 | 3 | 320,785 | 463,363,110 | 385,414 | 3,600,857 | 1* | -9 | -10 | -22 | 48142 | - |
|  | 10 | 4 | 70 | 82 | 3 | 214,632 | 620,130,770 | 278,324 | 2,922,624 | 1* | -39 | 21 | -37 | 41424 | - |
|  | 10 | 5 | 45 | 80 | 7 | 567,502 | 1,377,043,855 | 655,746 | 6,461,558 | 0* | 62 | 169 | 39 | 63939 | - |
|  | 10 | 5 | 45 | 75 | 6 | 554,118 | 1,254,714,856 | 635,199 | 6,148,904 | 0* | 58 | 145 | 33 | 59754 | - |
|  | 10 | 5 | 50 | 80 | 6 | 540,902 | 1,032,069,961 | 626,544 | 5,752,420 | 0* | 54 | 102 | 24 | 63240 | - |
|  | 10 | 5 | 50 | 75 | 5 | 526,696 | 904,318,681 | 604,872 | 5,421,222 | 0* | 50 | 77 | 17 | 58823 | - |
| Policy Position | Cut off (µg Hb/g) | Interval (Years) | Start Age | Stop Age | Number of Lifetime Screens | ΔQALY | ΔC (€) | ΔLYG | Total Number of Colonoscopies | Within Capacity 0= No 1= Yes | % ΔQALY | % ΔC | %ΔNo. Colonoscopies |  |  |
|  | 10 | 5 | 45 | 70 | 5 | 520,726 | 1,142,045,189 | 592,025 | 5,703,863 | 0* | 48 | 123 | 23 | 52903 | - |
|  | 10 | 5 | 50 | 70 | 4 | 490,702 | 786,836,047 | 558,424 | 4,950,911 | 0* | 40 | 54 | 7 | 51481 | - |
|  | 10 | 5 | 55 | 80 | 5 | 487,965 | 773,449,220 | 569,219 | 4,900,614 | 0* | 39 | 51 | 6 | 60535 | - |
| Max NHB within capacity (B) | 10 | 5 | 55 | 75 | 4 | 472,789 | 638,614,456 | 546,134 | 4,549,020 | 1* | 35 | 25 | -2 | 55821 | - |
|  | 10 | 5 | 45 | 65 | 4 | 463,521 | 1,052,653,478 | 523,459 | 5,110,920 | 0* | 32 | 106 | 10 | 44059 | - |
|  | 10 | 5 | 55 | 70 | 3 | 434,759 | 515,284,785 | 497,073 | 4,045,291 | 1* | 24 | 1 | -13 | 48018 | - |
|  | 10 | 5 | 50 | 65 | 3 | 430,174 | 692,863,637 | 485,786 | 4,316,919 | 1* | 22 | 35 | -7 | 42091 | - |
|  | 10 | 5 | 60 | 80 | 4 | 403,866 | 618,230,470 | 479,923 | 3,991,031 | 1* | 15 | 21 | -14 | 55257 | - |
|  | 10 | 5 | 60 | 75 | 3 | 387,639 | 475,577,032 | 455,246 | 3,611,471 | 1* | 10 | -7 | -22 | 50212 | - |
|  | 10 | 5 | 55 | 65 | 2 | 369,220 | 419,017,019 | 418,820 | 3,355,957 | 1* | 5 | -18 | -28 | 37875 | - |
|  | 10 | 5 | 45 | 60 | 3 | 379,983 | 974,232,295 | 428,176 | 4,348,822 | 1* | 8 | 90 | -6 | 33938 | - |
|  | 10 | 5 | 60 | 70 | 2 | 345,527 | 343,881,706 | 401,089 | 3,055,984 | 1* | -2 | -33 | -34 | 41599 | - |
|  | 10 | 5 | 50 | 60 | 2 | 340,757 | 610,099,492 | 383,614 | 3,492,848 | 1* | -3 | 19 | -25 | 31208 | - |
|  | 10 | 5 | 65 | 80 | 3 | 298,932 | 551,237,692 | 367,609 | 3,067,574 | 1* | -15 | 8 | -34 | 47215 | - |
|  | 10 | 5 | 65 | 75 | 2 | 280,903 | 395,944,154 | 340,307 | 2,649,399 | 1* | -20 | -23 | -43 | 41626 | - |
|  | 10 | 5 | 45 | 55 | 2 | 277,658 | 860,510,920 | 313,313 | 3,400,608 | 1* | -21 | 68 | -27 | 23507 | - |
|  | 10 | 5 | 70 | 80 | 2 | 193,019 | 539,979,484 | 252,102 | 2,185,244 | 1* | -45 | 5 | -53 | 36944 | - |
|  | 15 | 1 | 50 | 80 | 30 | 885,609 | 2,150,147,793 | 953,719 | 33,945,738 | 0 | 152 | 320 | 632 | 91214 | - |
|  | 15 | 1 | 50 | 75 | 25 | 867,906 | 2,008,183,578 | 933,175 | 33,209,233 | 0 | 147 | 292 | 616 | 87381 | - |
|  | 15 | 1 | 45 | 80 | 35 | 909,275 | 3,096,192,914 | 981,570 | 36,997,989 | 0 | 159 | 505 | 698 | 89670 | - |
|  | 15 | 1 | 45 | 75 | 30 | 893,801 | 2,975,861,303 | 963,697 | 36,367,643 | 0 | 154 | 481 | 684 | 86358 | - |
|  | 15 | 1 | 55 | 80 | 25 | 806,697 | 1,454,368,293 | 871,536 | 29,286,716 | 0 | 130 | 184 | 532 | 88908 | - |
|  | 15 | 1 | 50 | 70 | 20 | 825,582 | 1,839,383,141 | 886,891 | 31,474,667 | 0 | 135 | 259 | 579 | 80478 | - |
|  | 15 | 1 | 55 | 75 | 20 | 785,530 | 1,290,065,772 | 847,062 | 28,422,491 | 0 | 124 | 152 | 513 | 84364 | - |
|  | 15 | 1 | 45 | 70 | 25 | 858,183 | 2,828,337,696 | 924,693 | 34,884,576 | 0 | 144 | 452 | 652 | 80529 | - |
|  | 15 | 1 | 55 | 70 | 15 | 735,743 | 1,092,206,037 | 792,666 | 26,337,881 | 0 | 109 | 113 | 468 | 76246 | - |
|  | 15 | 1 | 50 | 65 | 15 | 744,930 | 1,635,877,295 | 801,275 | 28,269,083 | 0 | 112 | 219 | 510 | 69977 | - |
|  | 15 | 1 | 45 | 65 | 20 | 789,294 | 2,652,234,312 | 851,586 | 32,203,284 | 0 | 125 | 418 | 594 | 71565 | - |
|  | 15 | 1 | 60 | 80 | 20 | 661,581 | 1,030,873,996 | 725,098 | 23,405,975 | 0 | 88 | 101 | 405 | 81205 | - |
|  | 15 | 1 | 60 | 75 | 15 | 636,347 | 838,312,463 | 695,923 | 22,365,533 | 0 | 81 | 64 | 382 | 75775 | - |
|  | 15 | 1 | 55 | 65 | 10 | 636,977 | 863,045,712 | 688,211 | 22,370,084 | 0 | 81 | 69 | 382 | 63363 | - |
| Policy Position | Cut off (µg Hb/g) | Interval (Years) | Start Age | Stop Age | Number of Lifetime Screens | ΔQALY | ΔC (€) | ΔLYG | Total Number of Colonoscopies | Within Capacity 0= No 1= Yes | % ΔQALY | % ΔC | %ΔNo. Colonoscopies |  |  |
|  | 15 | 1 | 45 | 60 | 15 | 676,617 | 2,406,521,360 | 733,203 | 27,929,697 | 0 | 93 | 370 | 502 | 59346 | - |
|  | 15 | 1 | 60 | 70 | 10 | 575,369 | 611,537,554 | 629,627 | 19,751,682 | 0 | 64 | 19 | 326 | 65789 | - |
|  | 15 | 1 | 50 | 60 | 10 | 608,386 | 1,362,001,589 | 658,568 | 23,056,080 | 0 | 73 | 166 | 397 | 55258 | - |
|  | 15 | 1 | 65 | 80 | 15 | 485,781 | 819,627,956 | 547,921 | 16,859,864 | 0 | 38 | 60 | 264 | 69125 | - |
|  | 15 | 1 | 65 | 75 | 10 | 454,262 | 594,450,072 | 511,730 | 15,551,819 | 0 | 29 | 16 | 235 | 62400 | - |
|  | 15 | 1 | 45 | 55 | 10 | 514,212 | 2,022,129,051 | 562,876 | 21,745,446 | 0 | 46 | 295 | 369 | 43977 | - |
|  | 15 | 1 | 70 | 80 | 10 | 308,442 | 738,697,957 | 367,287 | 10,255,080 | 0 | -12 | 44 | 121 | 53668 | - |
|  | 15 | 2 | 45 | 81 | 18 | 769,895 | 2,057,488,235 | 866,988 | 16,469,886 | 0 | 119 | 302 | 255 | 82096 | - |
|  | 15 | 2 | 50 | 80 | 15 | 735,548 | 1,444,113,899 | 828,418 | 14,659,237 | 0 | 109 | 182 | 216 | 80981 | - |
|  | 15 | 2 | 50 | 76 | 13 | 720,404 | 1,331,638,049 | 808,151 | 14,272,441 | 0 | 105 | 160 | 208 | 77073 | - |
|  | 15 | 2 | 45 | 75 | 15 | 748,095 | 1,906,034,325 | 838,300 | 15,923,283 | 0 | 113 | 272 | 243 | 76607 | - |
|  | 15 | 2 | 45 | 71 | 13 | 714,781 | 1,805,924,149 | 798,669 | 15,240,898 | 0 | 103 | 253 | 229 | 70577 | - |
|  | 15 | 2 | 50 | 70 | 10 | 666,327 | 1,162,201,939 | 743,307 | 13,144,397 | 0 | 90 | 127 | 183 | 67312 | - |
|  | 15 | 2 | 55 | 81 | 13 | 659,853 | 1,041,042,150 | 748,216 | 12,485,887 | 0 | 88 | 103 | 169 | 78057 | - |
|  | 15 | 2 | 55 | 75 | 10 | 634,338 | 861,730,404 | 714,472 | 11,826,790 | 0 | 81 | 68 | 155 | 71631 | - |
|  | 15 | 2 | 55 | 71 | 8 | 594,817 | 741,262,291 | 667,374 | 10,988,441 | 0 | 69 | 45 | 137 | 64400 | - |
|  | 15 | 2 | 50 | 66 | 8 | 604,039 | 1,056,495,208 | 672,376 | 11,907,407 | 0 | 72 | 106 | 157 | 58434 | - |
|  | 15 | 2 | 45 | 65 | 10 | 628,902 | 1,653,603,954 | 700,933 | 13,568,232 | 0 | 79 | 223 | 193 | 58487 | - |
|  | 15 | 2 | 60 | 80 | 10 | 536,426 | 758,965,359 | 618,290 | 9,762,195 | 0 | 53 | 48 | 111 | 70055 | - |
|  | 15 | 2 | 60 | 76 | 8 | 517,214 | 629,615,563 | 593,155 | 9,284,595 | 0 | 47 | 23 | 100 | 65221 | - |
|  | 15 | 2 | 45 | 61 | 8 | 543,137 | 1,537,128,469 | 606,097 | 11,945,313 | 0 | 55 | 200 | 158 | 48672 | - |
|  | 15 | 2 | 55 | 65 | 5 | 487,200 | 574,426,306 | 545,444 | 8,849,283 | 0 | 39 | 12 | 91 | 49260 | - |
|  | 15 | 2 | 60 | 70 | 5 | 448,983 | 437,705,016 | 512,184 | 7,818,209 | 0 | 28 | -15 | 69 | 52886 | - |
|  | 15 | 2 | 50 | 60 | 5 | 457,516 | 879,019,251 | 511,038 | 9,151,145 | 0 | 30 | 72 | 97 | 41763 | - |
|  | 15 | 2 | 65 | 81 | 8 | 394,528 | 679,425,721 | 470,680 | 7,203,415 | 0* | 12 | 33 | 55 | 60278 | - |
|  | 15 | 2 | 65 | 75 | 5 | 360,621 | 467,362,112 | 426,770 | 6,333,236 | 0* | 3 | -9 | 37 | 51867 | - |
|  | 15 | 2 | 70 | 80 | 5 | 246,441 | 621,060,326 | 311,652 | 4,408,740 | 1* | -30 | 21 | -5 | 45548 | - |
|  | 15 | 3 | 45 | 81 | 12 | 654,996 | 1,668,914,859 | 758,141 | 9,672,427 | 0 | 86 | 226 | 109 | 72921 | - |
|  | 15 | 3 | 50 | 80 | 10 | 622,256 | 1,215,320,565 | 721,865 | 8,633,988 | 0 | 77 | 137 | 86 | 71484 | - |
|  | 15 | 3 | 45 | 75 | 10 | 634,516 | 1,511,280,937 | 728,699 | 9,238,874 | 0 | 81 | 195 | 99 | 67204 | - |
|  | 15 | 3 | 50 | 74 | 8 | 595,642 | 1,048,632,964 | 684,959 | 8,122,393 | 0 | 70 | 105 | 75 | 64642 | - |
| Policy Position | Cut off (µg Hb/g) | Interval (Years) | Start Age | Stop Age | Number of Lifetime Screens | ΔQALY | ΔC (€) | ΔLYG | Total Number of Colonoscopies | Within Capacity 0= No 1= Yes | % ΔQALY | % ΔC | %ΔNo. Colonoscopies |  |  |
|  | 15 | 3 | 50 | 71 | 7 | 569,044 | 970,156,138 | 651,535 | 7,726,530 | 0 | 62 | 89 | 67 | 59587 | - |
|  | 15 | 3 | 45 | 69 | 8 | 581,541 | 1,365,423,262 | 662,702 | 8,491,548 | 0 | 66 | 167 | 83 | 57518 | - |
|  | 15 | 3 | 55 | 79 | 8 | 552,602 | 863,361,916 | 644,772 | 7,287,037 | 0* | 57 | 69 | 57 | 67280 | - |
|  | 15 | 3 | 55 | 76 | 7 | 540,168 | 773,172,576 | 627,024 | 7,025,457 | 0* | 54 | 51 | 52 | 63851 | - |
|  | 15 | 3 | 45 | 66 | 7 | 539,799 | 1,301,237,031 | 613,332 | 7,958,937 | 0 | 54 | 154 | 72 | 51482 | - |
|  | 15 | 3 | 55 | 70 | 5 | 486,313 | 608,400,875 | 559,145 | 6,175,313 | 0* | 38 | 19 | 33 | 53338 | - |
|  | 15 | 3 | 50 | 65 | 5 | 483,671 | 833,700,767 | 550,290 | 6,576,703 | 0* | 38 | 63 | 42 | 46919 | - |
|  | 15 | 3 | 60 | 81 | 7 | 455,233 | 725,558,064 | 542,265 | 5,754,683 | 0* | 30 | 42 | 24 | 62293 | - |
|  | 15 | 3 | 60 | 75 | 5 | 429,959 | 538,913,629 | 506,230 | 5,213,198 | 0* | 22 | 5 | 12 | 55299 | - |
|  | 15 | 3 | 45 | 60 | 5 | 424,329 | 1,165,742,248 | 481,554 | 6,528,285 | 0* | 21 | 128 | 41 | 37810 | - |
|  | 15 | 3 | 60 | 69 | 3 | 360,768 | 372,266,673 | 421,118 | 4,217,763 | 1* | 3 | -27 | -9 | 42697 | - |
|  | 15 | 3 | 50 | 59 | 3 | 349,544 | 687,213,972 | 398,164 | 4,895,446 | 0* | -1 | 34 | 6 | 31661 | - |
|  | 15 | 3 | 65 | 80 | 5 | 330,490 | 611,984,366 | 407,183 | 4,217,313 | 1* | -6 | 20 | -9 | 51943 | - |
|  | 15 | 3 | 65 | 74 | 3 | 294,840 | 410,584,241 | 358,689 | 3,531,481 | 1* | -16 | -20 | -24 | 42955 | - |
|  | 15 | 3 | 45 | 54 | 3 | 277,111 | 958,068,240 | 316,672 | 4,626,028 | 1* | -21 | 87 | 0 | 23437 | - |
|  | 15 | 3 | 70 | 79 | 3 | 205,265 | 553,679,707 | 268,574 | 2,785,162 | 1* | -42 | 8 | -40 | 38809 | - |
|  | 15 | 4 | 45 | 81 | 9 | 565,662 | 1,471,592,383 | 668,389 | 6,610,138 | 0* | 61 | 187 | 43 | 65037 | - |
|  | 15 | 4 | 45 | 77 | 8 | 557,114 | 1,363,847,459 | 653,778 | 6,382,786 | 0* | 59 | 166 | 38 | 61887 | - |
|  | 15 | 4 | 50 | 82 | 8 | 539,523 | 1,148,231,612 | 640,315 | 5,979,136 | 0* | 54 | 124 | 29 | 64797 | - |
|  | 15 | 4 | 50 | 74 | 6 | 514,302 | 930,422,368 | 601,613 | 5,461,626 | 0* | 46 | 82 | 18 | 57237 | - |
|  | 15 | 4 | 45 | 69 | 6 | 500,831 | 1,167,339,795 | 579,731 | 5,697,125 | 0* | 43 | 128 | 23 | 50403 | - |
|  | 15 | 4 | 55 | 79 | 6 | 480,199 | 803,947,912 | 571,221 | 5,018,591 | 0* | 37 | 57 | 8 | 60113 | - |
|  | 15 | 4 | 50 | 70 | 5 | 480,940 | 829,351,044 | 558,268 | 5,064,781 | 0* | 37 | 62 | 9 | 50717 | - |
|  | 15 | 4 | 55 | 75 | 5 | 463,990 | 687,933,364 | 547,314 | 4,716,996 | 0* | 32 | 34 | 2 | 55511 | - |
|  | 15 | 4 | 55 | 71 | 4 | 431,832 | 580,051,619 | 505,190 | 4,300,085 | 1* | 23 | 13 | -7 | 48887 | - |
|  | 15 | 4 | 45 | 65 | 5 | 449,429 | 1,088,037,055 | 517,452 | 5,190,493 | 0* | 28 | 113 | 12 | 42795 | - |
|  | 15 | 4 | 50 | 66 | 4 | 430,390 | 743,983,212 | 496,577 | 4,544,728 | 1* | 23 | 45 | -2 | 42868 | - |
|  | 15 | 4 | 60 | 80 | 5 | 409,263 | 716,240,120 | 495,611 | 4,262,037 | 1* | 16 | 40 | -8 | 57888 | - |
|  | 15 | 4 | 55 | 67 | 3 | 383,196 | 488,753,783 | 445,081 | 3,753,314 | 1* | 9 | -5 | -19 | 40854 | - |
|  | 15 | 4 | 60 | 76 | 4 | 381,041 | 542,421,244 | 457,915 | 3,668,694 | 1* | 8 | 6 | -21 | 50628 | - |
|  | 15 | 4 | 45 | 61 | 4 | 380,945 | 1,009,566,034 | 437,560 | 4,554,086 | 1* | 8 | 97 | -2 | 34476 | - |
| Policy Position | Cut off (µg Hb/g) | Interval (Years) | Start Age | Stop Age | Number of Lifetime Screens | ΔQALY | ΔC (€) | ΔLYG | Total Number of Colonoscopies | Within Capacity 0= No 1= Yes | % ΔQALY | % ΔC | %ΔNo. Colonoscopies |  |  |
|  | 15 | 4 | 60 | 72 | 3 | 350,936 | 427,059,057 | 417,828 | 3,268,939 | 1* | 0 | -17 | -30 | 44028 | - |
|  | 15 | 4 | 50 | 62 | 3 | 361,793 | 665,847,270 | 416,182 | 3,888,870 | 1* | 3 | 30 | -16 | 34122 | - |
|  | 15 | 4 | 65 | 81 | 4 | 290,888 | 623,243,466 | 367,405 | 2,992,424 | 1* | -17 | 22 | -35 | 47367 | - |
|  | 15 | 4 | 65 | 77 | 3 | 279,325 | 493,845,071 | 348,431 | 2,696,038 | 1* | -20 | -4 | -42 | 43293 | - |
|  | 15 | 4 | 45 | 57 | 3 | 300,105 | 914,686,502 | 344,989 | 3,789,925 | 1* | -15 | 79 | -18 | 25950 | - |
|  | 15 | 4 | 70 | 82 | 3 | 188,010 | 629,273,922 | 254,155 | 2,207,244 | 1* | -46 | 23 | -52 | 37695 | - |
|  | 15 | 5 | 45 | 80 | 7 | 500,610 | 1,313,850,021 | 598,861 | 4,935,619 | 0* | 42 | 157 | 6 | 58544 | - |
|  | 15 | 5 | 45 | 75 | 6 | 486,677 | 1,180,942,028 | 576,383 | 4,673,051 | 0* | 39 | 131 | 1 | 54020 | - |
|  | 15 | 5 | 50 | 80 | 6 | 475,679 | 1,015,483,005 | 570,519 | 4,357,665 | 1* | 35 | 98 | -6 | 57598 | - |
|  | 15 | 5 | 50 | 75 | 5 | 461,277 | 878,718,250 | 547,342 | 4,083,026 | 1* | 31 | 72 | -12 | 52912 | - |
|  | 15 | 5 | 45 | 70 | 5 | 452,904 | 1,060,740,744 | 531,053 | 4,315,689 | 1* | 29 | 107 | -7 | 46903 | - |
|  | 15 | 5 | 50 | 70 | 4 | 425,468 | 755,918,185 | 499,406 | 3,709,683 | 1* | 21 | 48 | -20 | 45441 | - |
|  | 15 | 5 | 55 | 80 | 5 | 427,034 | 791,205,827 | 516,150 | 3,678,713 | 1* | 22 | 55 | -21 | 54882 | - |
|  | 15 | 5 | 55 | 75 | 4 | 411,822 | 648,100,598 | 491,754 | 3,390,299 | 1* | 17 | 27 | -27 | 49954 | - |
|  | 15 | 5 | 45 | 65 | 4 | 397,544 | 963,129,558 | 462,149 | 3,850,646 | 1* | 13 | 88 | -17 | 38189 | - |
|  | 15 | 5 | 55 | 70 | 3 | 374,489 | 519,908,398 | 441,748 | 2,996,199 | 1* | 7 | 2 | -35 | 42111 | - |
|  | 15 | 5 | 50 | 65 | 3 | 368,194 | 654,927,801 | 428,023 | 3,220,490 | 1* | 5 | 28 | -31 | 36373 | - |
|  | 15 | 5 | 60 | 80 | 4 | 351,784 | 648,620,717 | 433,826 | 2,968,500 | 1* | 0 | 27 | -36 | 49887 | - |
|  | 15 | 5 | 60 | 75 | 3 | 335,822 | 500,117,407 | 408,271 | 2,661,968 | 1* | -4 | -2 | -43 | 44712 | - |
|  | 15 | 5 | 55 | 65 | 2 | 313,534 | 416,472,620 | 366,077 | 2,467,491 | 1* | -11 | -19 | -47 | 32476 | - |
|  | 15 | 5 | 60 | 70 | 2 | 295,579 | 363,502,321 | 354,389 | 2,231,746 | 1* | -16 | -29 | -52 | 36253 | - |
|  | 15 | 5 | 45 | 60 | 3 | 320,413 | 876,017,339 | 371,037 | 3,261,336 | 1* | -9 | 71 | -30 | 28709 | - |
|  | 15 | 5 | 50 | 60 | 2 | 286,035 | 566,190,578 | 331,010 | 2,592,360 | 1* | -19 | 11 | -44 | 26281 | - |
|  | 15 | 5 | 65 | 80 | 3 | 260,180 | 573,602,400 | 332,360 | 2,262,391 | 1* | -26 | 12 | -51 | 42537 | - |
|  | 15 | 5 | 65 | 75 | 2 | 242,585 | 414,119,308 | 304,378 | 1,928,911 | 1* | -31 | -19 | -58 | 36913 | - |
|  | 15 | 5 | 45 | 55 | 2 | 228,575 | 762,681,838 | 265,001 | 2,533,203 | 1* | -35 | 49 | -45 | 19317 | - |
|  | 15 | 5 | 70 | 80 | 2 | 167,463 | 544,126,708 | 227,634 | 1,604,568 | 1* | -52 | 6 | -65 | 33141 | - |
|  | 20 | 1 | 50 | 80 | 30 | 852,468 | 2,137,772,800 | 936,679 | 31,270,289 | 0 | 143 | 318 | 574 | 90065 | - |
|  | 20 | 1 | 45 | 80 | 35 | 885,735 | 3,034,168,533 | 974,716 | 34,532,939 | 0 | 152 | 493 | 645 | 89853 | - |
|  | 20 | 1 | 50 | 75 | 25 | 831,254 | 1,977,549,582 | 911,352 | 30,529,828 | 0 | 137 | 286 | 558 | 85376 | - |
|  | 20 | 1 | 45 | 75 | 30 | 866,195 | 2,892,364,773 | 951,525 | 33,874,407 | 0 | 147 | 465 | 630 | 85600 | - |
| Policy Position | Cut off (µg Hb/g) | Interval (Years) | Start Age | Stop Age | Number of Lifetime Screens | ΔQALY | ΔC (€) | ΔLYG | Total Number of Colonoscopies | Within Capacity 0= No 1= Yes | % ΔQALY | % ΔC | %ΔNo. Colonoscopies |  |  |
|  | 20 | 1 | 55 | 80 | 25 | 770,114 | 1,463,537,276 | 848,658 | 26,629,041 | 0 | 119 | 186 | 474 | 86886 | - |
|  | 20 | 1 | 50 | 70 | 20 | 783,162 | 1,796,565,424 | 857,576 | 28,791,947 | 0 | 123 | 251 | 521 | 77425 | - |
|  | 20 | 1 | 45 | 70 | 25 | 823,919 | 2,728,765,832 | 904,221 | 32,336,009 | 0 | 135 | 433 | 597 | 78605 | - |
|  | 20 | 1 | 55 | 75 | 20 | 745,519 | 1,285,609,223 | 819,447 | 25,791,141 | 0 | 112 | 151 | 456 | 81503 | - |
|  | 20 | 1 | 55 | 70 | 15 | 690,314 | 1,082,003,696 | 757,769 | 23,771,829 | 0 | 96 | 111 | 413 | 72395 | - |
|  | 20 | 1 | 45 | 65 | 20 | 747,052 | 2,538,769,756 | 820,740 | 29,602,790 | 0 | 113 | 396 | 538 | 68491 | - |
|  | 20 | 1 | 50 | 65 | 15 | 696,137 | 1,586,715,984 | 763,108 | 25,638,880 | 0 | 98 | 210 | 453 | 65967 | - |
|  | 20 | 1 | 60 | 80 | 20 | 628,827 | 1,046,992,635 | 702,983 | 21,013,052 | 0 | 79 | 104 | 353 | 78830 | - |
|  | 20 | 1 | 60 | 75 | 15 | 600,425 | 845,461,430 | 669,287 | 20,034,408 | 0 | 71 | 65 | 332 | 72598 | - |
|  | 20 | 1 | 55 | 65 | 10 | 587,417 | 854,814,872 | 646,616 | 19,986,146 | 0 | 67 | 67 | 331 | 58873 | - |
|  | 20 | 1 | 45 | 60 | 15 | 626,993 | 2,286,176,874 | 692,304 | 25,371,453 | 0 | 78 | 347 | 447 | 55449 | - |
|  | 20 | 1 | 60 | 70 | 10 | 535,732 | 620,132,854 | 597,265 | 17,574,762 | 0 | 52 | 21 | 279 | 61865 | - |
|  | 20 | 1 | 50 | 60 | 10 | 556,406 | 1,310,091,119 | 613,898 | 20,631,714 | 0 | 58 | 156 | 345 | 50837 | - |
|  | 20 | 1 | 65 | 80 | 15 | 459,805 | 824,824,424 | 528,854 | 14,907,893 | 0 | 31 | 61 | 221 | 66734 | - |
|  | 20 | 1 | 65 | 75 | 10 | 425,413 | 598,496,206 | 488,396 | 13,709,428 | 0 | 21 | 17 | 196 | 59277 | - |
|  | 20 | 1 | 45 | 55 | 10 | 463,401 | 1,898,756,805 | 517,279 | 19,430,262 | 0 | 32 | 271 | 319 | 39932 | - |
|  | 20 | 1 | 70 | 80 | 10 | 290,429 | 732,131,472 | 352,853 | 8,897,294 | 0 | -17 | 43 | 92 | 51494 | - |
|  | 20 | 2 | 45 | 81 | 18 | 724,960 | 1,988,589,847 | 831,857 | 14,389,148 | 0 | 106 | 288 | 210 | 79212 | - |
|  | 20 | 2 | 50 | 80 | 15 | 688,245 | 1,421,745,756 | 790,096 | 12,688,793 | 0 | 96 | 178 | 174 | 77462 | - |
|  | 20 | 2 | 45 | 75 | 15 | 701,142 | 1,822,288,770 | 799,504 | 13,880,229 | 0 | 100 | 256 | 199 | 73080 | - |
|  | 20 | 2 | 50 | 76 | 13 | 671,671 | 1,301,914,981 | 767,360 | 12,337,457 | 0 | 91 | 154 | 166 | 73122 | - |
|  | 20 | 2 | 45 | 71 | 13 | 665,798 | 1,714,704,364 | 756,367 | 13,247,347 | 0 | 90 | 235 | 186 | 66590 | - |
|  | 20 | 2 | 55 | 81 | 13 | 614,810 | 1,045,720,282 | 710,900 | 10,765,996 | 0 | 75 | 104 | 132 | 74397 | - |
|  | 20 | 2 | 50 | 70 | 10 | 615,550 | 1,124,458,783 | 698,461 | 11,314,090 | 0 | 75 | 120 | 144 | 62797 | - |
| Initial recommendation Policy (1) | 20 | 2 | 55 | 75 | 10 | 587,514 | 857,478,098 | 673,872 | 10,169,898 | 0 | 67 | 67 | 119 | 67404 | - |
|  | 20 | 2 | 55 | 71 | 8 | 547,167 | 732,966,124 | 624,463 | 9,416,543 | 0 | 56 | 43 | 103 | 59881 | - |
|  | 20 | 2 | 50 | 66 | 8 | 552,972 | 1,016,800,276 | 625,753 | 10,204,053 | 0 | 57 | 99 | 120 | 53825 | - |
|  | 20 | 2 | 45 | 65 | 10 | 577,236 | 1,557,853,051 | 653,501 | 11,711,980 | 0 | 64 | 204 | 153 | 53969 | - |
|  | 20 | 2 | 60 | 80 | 10 | 498,556 | 769,646,702 | 585,738 | 8,330,943 | 0 | 42 | 50 | 80 | 66450 | - |
|  | 20 | 2 | 60 | 76 | 8 | 478,611 | 635,781,678 | 558,864 | 7,909,959 | 0 | 36 | 24 | 71 | 61314 | - |
|  | 20 | 2 | 45 | 61 | 8 | 492,305 | 1,439,698,235 | 557,923 | 10,246,097 | 0 | 40 | 181 | 121 | 44231 | - |
| Policy Position | Cut off (µg Hb/g) | Interval (Years) | Start Age | Stop Age | Number of Lifetime Screens | ΔQALY | ΔC (€) | ΔLYG | Total Number of Colonoscopies | Within Capacity 0= No 1= Yes | % ΔQALY | % ΔC | %ΔNo. Colonoscopies |  |  |
|  | 20 | 2 | 55 | 65 | 5 | 441,059 | 566,241,695 | 501,764 | 7,522,059 | 0 | 26 | 11 | 62 | 44824 | - |
| Age Restriction (2) | 20 | 2 | 60 | 70 | 5 | 409,868 | 444,216,122 | 475,514 | 6,624,968 | 0* | 17 | -13 | 43 | 48722 | - |
|  | 20 | 2 | 50 | 60 | 5 | 409,843 | 837,441,676 | 465,066 | 7,757,511 | 0 | 17 | 64 | 67 | 37442 | - |
|  | 20 | 2 | 65 | 81 | 8 | 366,110 | 691,292,756 | 445,665 | 6,114,863 | 0* | 4 | 35 | 32 | 57065 | - |
|  | 20 | 2 | 65 | 75 | 5 | 331,474 | 474,925,597 | 399,499 | 5,353,641 | 0* | -6 | -7 | 15 | 48311 | - |
|  | 20 | 2 | 45 | 55 | 5 | 331,722 | 1,182,725,297 | 378,856 | 7,378,623 | 0 | -6 | 131 | 59 | 28389 | - |
|  | 20 | 2 | 70 | 80 | 5 | 227,516 | 620,666,855 | 293,741 | 3,680,243 | 1* | -35 | 21 | -21 | 42862 | - |
|  | 20 | 3 | 45 | 81 | 12 | 607,692 | 1,602,741,230 | 715,814 | 8,191,639 | 0 | 73 | 213 | 77 | 69124 | - |
|  | 20 | 3 | 45 | 75 | 10 | 586,140 | 1,435,413,852 | 684,020 | 7,802,609 | 0 | 67 | 180 | 68 | 63012 | - |
|  | 20 | 3 | 50 | 80 | 10 | 573,034 | 1,187,605,873 | 677,014 | 7,245,190 | 0* | 63 | 132 | 56 | 67280 | - |
|  | 20 | 3 | 50 | 74 | 8 | 545,705 | 1,012,911,190 | 638,103 | 6,797,736 | 0* | 55 | 98 | 47 | 60130 | - |
|  | 20 | 3 | 50 | 71 | 7 | 518,994 | 932,863,224 | 603,854 | 6,455,159 | 0* | 48 | 82 | 39 | 54974 | - |
|  | 20 | 3 | 45 | 69 | 8 | 532,373 | 1,284,455,158 | 615,734 | 7,145,554 | 0* | 52 | 151 | 54 | 53061 | - |
|  | 20 | 3 | 55 | 79 | 8 | 507,917 | 860,102,287 | 603,449 | 6,116,034 | 0* | 45 | 68 | 32 | 62996 | - |
|  | 20 | 3 | 55 | 76 | 7 | 495,267 | 765,764,419 | 584,838 | 5,886,280 | 0* | 41 | 50 | 27 | 59427 | - |
|  | 20 | 3 | 45 | 66 | 7 | 491,009 | 1,218,944,942 | 565,913 | 6,679,975 | 0* | 40 | 138 | 44 | 47027 | - |
|  | 20 | 3 | 55 | 70 | 5 | 441,427 | 597,831,530 | 515,566 | 5,149,904 | 0* | 26 | 17 | 11 | 48819 | - |
|  | 20 | 3 | 50 | 65 | 5 | 435,109 | 794,212,550 | 502,525 | 5,467,080 | 0* | 24 | 55 | 18 | 42417 | - |
|  | 20 | 3 | 60 | 81 | 7 | 417,952 | 734,013,070 | 507,384 | 4,801,017 | 0* | 19 | 43 | 4 | 58364 | - |
|  | 20 | 3 | 60 | 75 | 5 | 392,721 | 542,333,555 | 470,338 | 4,332,590 | 1* | 12 | 6 | -7 | 51190 | - |
|  | 20 | 3 | 55 | 64 | 3 | 342,454 | 461,181,246 | 397,826 | 3,965,106 | 1* | -3 | -10 | -14 | 34751 | - |
|  | 20 | 3 | 60 | 69 | 3 | 325,039 | 373,273,737 | 385,321 | 3,483,854 | 1* | -7 | -27 | -25 | 38745 | - |
|  | 20 | 3 | 50 | 59 | 3 | 308,809 | 645,763,766 | 356,699 | 4,037,432 | 1* | -12 | 26 | -13 | 27960 | - |
|  | 20 | 3 | 65 | 80 | 5 | 302,956 | 615,758,970 | 380,449 | 3,476,636 | 1* | -14 | 20 | -25 | 48500 | - |
|  | 20 | 3 | 65 | 74 | 3 | 267,897 | 412,000,819 | 331,476 | 2,894,658 | 1* | -24 | -20 | -38 | 39468 | - |
|  | 20 | 3 | 45 | 54 | 3 | 242,022 | 876,139,858 | 280,040 | 3,813,237 | 1* | -31 | 71 | -18 | 20336 | - |
|  | 20 | 3 | 70 | 79 | 3 | 187,467 | 547,161,952 | 249,992 | 2,295,719 | 1* | -47 | 7 | -50 | 36030 | - |
|  | 20 | 4 | 45 | 81 | 9 | 518,792 | 1,402,355,729 | 623,252 | 5,478,181 | 0* | 48 | 174 | 18 | 60870 | - |
|  | 20 | 4 | 45 | 77 | 8 | 510,013 | 1,290,070,987 | 607,841 | 5,277,742 | 0* | 45 | 152 | 14 | 57568 | - |
|  | 20 | 4 | 50 | 82 | 8 | 492,892 | 1,114,880,456 | 595,250 | 4,951,755 | 0* | 40 | 118 | 7 | 60489 | - |
|  | 20 | 4 | 50 | 74 | 6 | 467,483 | 889,451,220 | 555,250 | 4,500,392 | 1* | 33 | 74 | -3 | 52741 | - |
| Policy Position | Cut off (µg Hb/g) | Interval (Years) | Start Age | Stop Age | Number of Lifetime Screens | ΔQALY | ΔC (€) | ΔLYG | Total Number of Colonoscopies | Within Capacity 0= No 1= Yes | % ΔQALY | % ΔC | %ΔNo. Colonoscopies |  |  |
|  | 20 | 4 | 45 | 69 | 6 | 454,086 | 1,087,994,374 | 532,793 | 4,690,172 | 0* | 29 | 112 | 1 | 45994 | - |
|  | 20 | 4 | 55 | 79 | 6 | 437,789 | 792,559,390 | 529,734 | 4,139,237 | 1* | 25 | 55 | -11 | 55772 | - |
|  | 20 | 4 | 50 | 70 | 5 | 434,544 | 786,324,715 | 511,679 | 4,162,676 | 1* | 24 | 54 | -10 | 46247 | - |
|  | 20 | 4 | 55 | 75 | 5 | 421,753 | 673,983,811 | 505,463 | 3,879,373 | 1* | 20 | 32 | -16 | 51120 | - |
|  | 20 | 4 | 55 | 71 | 4 | 390,248 | 564,980,505 | 463,329 | 3,525,942 | 1* | 11 | 10 | -24 | 44543 | - |
|  | 20 | 4 | 45 | 65 | 5 | 404,356 | 1,006,910,833 | 471,470 | 4,261,725 | 1* | 15 | 97 | -8 | 38577 | - |
|  | 20 | 4 | 50 | 66 | 4 | 385,926 | 700,220,442 | 451,316 | 3,725,365 | 1* | 10 | 37 | -20 | 38629 | - |
|  | 20 | 4 | 60 | 80 | 5 | 374,464 | 717,183,825 | 461,673 | 3,496,816 | 1* | 7 | 40 | -25 | 54025 | - |
|  | 20 | 4 | 55 | 67 | 3 | 343,670 | 473,498,944 | 404,714 | 3,066,955 | 1* | -2 | -8 | -34 | 36785 | - |
|  | 20 | 4 | 60 | 76 | 4 | 346,466 | 537,687,313 | 422,999 | 2,985,970 | 1* | -1 | 5 | -36 | 46645 | - |
|  | 20 | 4 | 60 | 72 | 3 | 317,008 | 422,078,644 | 383,054 | 2,651,791 | 1* | -10 | -18 | -43 | 40116 | - |
|  | 20 | 4 | 45 | 61 | 4 | 339,054 | 929,166,460 | 394,222 | 3,726,084 | 1* | -3 | 81 | -20 | 30612 | - |
|  | 20 | 4 | 50 | 62 | 3 | 321,181 | 622,274,818 | 374,224 | 3,175,748 | 1* | -9 | 22 | -32 | 30312 | - |
|  | 20 | 4 | 65 | 81 | 4 | 265,039 | 620,584,899 | 340,918 | 2,440,677 | 1* | -25 | 21 | -47 | 43923 | - |
|  | 20 | 4 | 65 | 77 | 3 | 253,707 | 490,041,966 | 321,785 | 2,188,537 | 1* | -28 | -4 | -53 | 39852 | - |
|  | 20 | 4 | 45 | 57 | 3 | 264,302 | 834,617,702 | 307,287 | 3,090,030 | 1* | -25 | 63 | -33 | 22711 | - |
|  | 20 | 4 | 70 | 82 | 3 | 170,983 | 617,294,374 | 235,626 | 1,804,350 | 1* | -51 | 21 | -61 | 34896 | - |
|  | 20 | 5 | 45 | 80 | 7 | 455,853 | 1,243,592,264 | 553,786 | 4,020,546 | 1* | 30 | 143 | -13 | 54291 | - |
|  | 20 | 5 | 45 | 75 | 6 | 441,807 | 1,108,012,569 | 530,714 | 3,795,396 | 1* | 26 | 116 | -18 | 49676 | - |
|  | 20 | 5 | 50 | 80 | 6 | 431,501 | 977,626,063 | 525,971 | 3,538,069 | 1* | 23 | 91 | -24 | 53195 | - |
|  | 20 | 5 | 50 | 75 | 5 | 417,301 | 837,930,527 | 502,525 | 3,303,644 | 1* | 19 | 64 | -29 | 48484 | - |
|  | 20 | 5 | 45 | 70 | 5 | 408,630 | 986,434,727 | 485,418 | 3,497,136 | 1* | 16 | 93 | -25 | 42635 | - |
|  | 20 | 5 | 55 | 80 | 5 | 387,642 | 773,856,182 | 476,197 | 2,975,623 | 1* | 10 | 51 | -36 | 50665 | - |
|  | 20 | 5 | 50 | 70 | 4 | 382,544 | 713,081,177 | 455,132 | 2,992,787 | 1* | 9 | 39 | -35 | 41164 | - |
|  | 20 | 5 | 55 | 75 | 4 | 372,872 | 628,937,138 | 451,911 | 2,731,046 | 1* | 6 | 23 | -41 | 45790 | - |
|  | 20 | 5 | 55 | 70 | 3 | 336,584 | 501,385,392 | 402,539 | 2,406,805 | 1* | -4 | -2 | -48 | 38104 | - |
|  | 20 | 5 | 45 | 65 | 4 | 355,563 | 887,926,065 | 418,301 | 3,114,082 | 1* | 1 | 73 | -33 | 34222 | - |
|  | 20 | 5 | 50 | 65 | 3 | 328,183 | 612,000,911 | 386,348 | 2,592,142 | 1* | -7 | 20 | -44 | 32496 | - |
|  | 20 | 5 | 60 | 80 | 4 | 319,227 | 638,731,511 | 400,067 | 2,392,408 | 1* | -9 | 25 | -48 | 46050 | - |
|  | 20 | 5 | 60 | 75 | 3 | 303,575 | 488,947,559 | 374,426 | 2,132,932 | 1* | -14 | -5 | -54 | 40899 | - |
|  | 20 | 5 | 55 | 65 | 2 | 279,206 | 397,977,075 | 330,092 | 1,976,801 | 1* | -21 | -22 | -57 | 28966 | - |
| Policy Position | Cut off (µg Hb/g) | Interval (Years) | Start Age | Stop Age | Number of Lifetime Screens | ΔQALY | ΔC (€) | ΔLYG | Total Number of Colonoscopies | Within Capacity 0= No 1= Yes | % ΔQALY | % ΔC | %ΔNo. Colonoscopies |  |  |
|  | 20 | 5 | 60 | 70 | 2 | 264,818 | 353,748,083 | 321,707 | 1,780,406 | 1* | -25 | -31 | -62 | 32688 | - |
|  | 20 | 5 | 45 | 60 | 3 | 283,754 | 799,465,759 | 332,116 | 2,630,256 | 1* | -19 | 56 | -43 | 25342 | - |
|  | 20 | 5 | 50 | 60 | 2 | 251,933 | 522,159,389 | 294,910 | 2,083,252 | 1* | -28 | 2 | -55 | 23117 | - |
|  | 20 | 5 | 65 | 80 | 3 | 235,818 | 562,593,335 | 306,231 | 1,813,703 | 1* | -33 | 10 | -61 | 39163 | - |
|  | 20 | 5 | 65 | 75 | 2 | 218,843 | 403,992,113 | 278,602 | 1,534,452 | 1* | -38 | -21 | -67 | 33645 | - |
|  | 20 | 5 | 45 | 55 | 2 | 199,724 | 689,374,791 | 233,929 | 2,033,822 | 1* | -43 | 35 | -56 | 16758 | - |
|  | 20 | 5 | 70 | 80 | 2 | 151,559 | 526,025,077 | 209,483 | 1,284,537 | 1* | -57 | 3 | -72 | 30427 | - |
|  | 30 | 1 | 50 | 80 | 30 | 821,429 | 2,156,508,902 | 924,037 | 28,007,745 | 0 | 134 | 321 | 504 | 88939 | - |
|  | 30 | 1 | 45 | 80 | 35 | 861,614 | 3,015,420,600 | 970,052 | 31,244,767 | 0 | 145 | 489 | 574 | 89680 | - |
|  | 30 | 1 | 50 | 75 | 25 | 796,496 | 1,981,667,521 | 893,397 | 27,296,663 | 0 | 127 | 287 | 489 | 83311 | - |
|  | 30 | 1 | 45 | 75 | 30 | 838,437 | 2,855,198,793 | 941,664 | 30,596,676 | 0 | 139 | 458 | 560 | 84505 | - |
|  | 30 | 1 | 55 | 80 | 25 | 737,501 | 1,497,042,428 | 831,821 | 23,638,436 | 0 | 110 | 192 | 410 | 85153 | - |
|  | 30 | 1 | 45 | 70 | 25 | 789,739 | 2,677,339,792 | 885,597 | 29,083,690 | 0 | 125 | 423 | 527 | 76368 | - |
|  | 30 | 1 | 50 | 70 | 20 | 743,143 | 1,789,477,510 | 832,059 | 25,625,686 | 0 | 112 | 249 | 453 | 74360 | - |
|  | 30 | 1 | 55 | 75 | 20 | 709,723 | 1,307,005,465 | 797,834 | 22,852,408 | 0 | 102 | 155 | 393 | 78904 | - |
|  | 30 | 1 | 55 | 70 | 15 | 649,986 | 1,096,698,020 | 729,250 | 20,961,469 | 0 | 85 | 114 | 352 | 68936 | - |
|  | 30 | 1 | 45 | 65 | 20 | 705,865 | 2,475,340,477 | 792,134 | 26,432,829 | 0 | 101 | 383 | 470 | 65249 | - |
|  | 30 | 1 | 50 | 65 | 15 | 650,563 | 1,570,617,520 | 728,818 | 22,649,850 | 0 | 85 | 207 | 388 | 62086 | - |
|  | 30 | 1 | 60 | 80 | 20 | 599,444 | 1,083,990,558 | 686,335 | 18,470,008 | 0 | 71 | 112 | 298 | 76802 | - |
|  | 30 | 1 | 60 | 75 | 15 | 568,154 | 874,202,124 | 648,058 | 17,575,158 | 0 | 62 | 71 | 279 | 69788 | - |
|  | 30 | 1 | 55 | 65 | 10 | 543,570 | 870,248,079 | 611,577 | 17,472,616 | 0 | 55 | 70 | 277 | 54879 | - |
|  | 30 | 1 | 45 | 60 | 15 | 581,588 | 2,211,379,444 | 656,010 | 22,427,299 | 0 | 66 | 332 | 384 | 51667 | - |
|  | 30 | 1 | 60 | 70 | 10 | 499,949 | 649,146,776 | 570,158 | 15,327,192 | 0 | 42 | 27 | 231 | 58353 | - |
|  | 30 | 1 | 50 | 60 | 10 | 510,105 | 1,288,055,672 | 575,166 | 18,033,736 | 0 | 45 | 152 | 289 | 46774 | - |
|  | 30 | 1 | 65 | 80 | 15 | 436,718 | 850,829,582 | 514,628 | 12,959,958 | 0 | 24 | 66 | 179 | 64738 | - |
|  | 30 | 1 | 65 | 75 | 10 | 399,869 | 622,268,209 | 470,060 | 11,886,997 | 0 | 14 | 22 | 156 | 56598 | - |
|  | 30 | 1 | 45 | 55 | 10 | 419,789 | 1,816,904,748 | 478,951 | 16,960,210 | 0 | 19 | 255 | 266 | 36281 | - |
|  | 30 | 1 | 70 | 80 | 10 | 274,335 | 745,348,669 | 342,071 | 7,626,031 | 0 | -22 | 46 | 64 | 49674 | - |
|  | 30 | 2 | 45 | 81 | 18 | 690,163 | 1,962,236,298 | 811,561 | 12,336,037 | 0 | 96 | 283 | 166 | 77354 | - |
|  | 30 | 2 | 50 | 80 | 15 | 650,493 | 1,429,284,981 | 765,424 | 10,781,262 | 0 | 85 | 179 | 132 | 75012 | - |
|  | 30 | 2 | 45 | 75 | 15 | 664,462 | 1,780,535,838 | 775,328 | 11,875,157 | 0 | 89 | 248 | 156 | 70539 | - |
| Policy Position | Cut off (µg Hb/g) | Interval (Years) | Start Age | Stop Age | Number of Lifetime Screens | ΔQALY | ΔC (€) | ΔLYG | Total Number of Colonoscopies | Within Capacity 0= No 1= Yes | % ΔQALY | % ΔC | %ΔNo. Colonoscopies |  |  |
|  | 30 | 2 | 50 | 76 | 13 | 632,610 | 1,301,083,286 | 740,122 | 10,470,691 | 0 | 80 | 154 | 126 | 70243 | - |
|  | 30 | 2 | 45 | 71 | 13 | 627,306 | 1,665,398,540 | 728,625 | 11,305,367 | 0 | 79 | 225 | 144 | 63593 | - |
|  | 30 | 2 | 55 | 81 | 13 | 580,183 | 1,077,242,242 | 688,198 | 9,130,679 | 0 | 65 | 110 | 97 | 71934 | - |
|  | 30 | 2 | 50 | 70 | 10 | 574,408 | 1,115,559,506 | 666,631 | 9,572,835 | 0 | 64 | 118 | 106 | 59310 | - |
|  | 30 | 2 | 55 | 75 | 10 | 551,440 | 879,134,937 | 647,906 | 8,605,265 | 0 | 57 | 72 | 86 | 64381 | - |
|  | 30 | 2 | 55 | 71 | 8 | 509,958 | 750,754,394 | 595,739 | 7,942,711 | 0 | 45 | 47 | 71 | 56544 | - |
|  | 30 | 2 | 45 | 65 | 10 | 536,485 | 1,500,552,285 | 620,361 | 9,942,069 | 0 | 53 | 193 | 114 | 50523 | - |
|  | 30 | 2 | 50 | 66 | 8 | 511,332 | 1,003,830,729 | 591,408 | 8,602,124 | 0 | 46 | 96 | 86 | 50152 | - |
|  | 30 | 2 | 60 | 80 | 10 | 468,365 | 806,004,709 | 564,866 | 6,991,978 | 0* | 33 | 57 | 51 | 63960 | - |
|  | 30 | 2 | 60 | 76 | 8 | 447,648 | 666,088,528 | 535,990 | 6,629,173 | 0* | 27 | 30 | 43 | 58484 | - |
|  | 30 | 2 | 45 | 61 | 8 | 452,061 | 1,376,256,330 | 522,945 | 8,654,539 | 0 | 29 | 169 | 87 | 40752 | - |
|  | 30 | 2 | 55 | 65 | 5 | 404,956 | 578,532,013 | 471,177 | 6,302,288 | 0* | 15 | 13 | 36 | 41462 | - |
|  | 30 | 2 | 50 | 60 | 5 | 372,630 | 816,978,413 | 431,851 | 6,488,748 | 0* | 6 | 60 | 40 | 34105 | - |
|  | 30 | 2 | 65 | 81 | 8 | 342,912 | 723,060,020 | 429,076 | 5,128,835 | 0* | -2 | 41 | 11 | 54791 | - |
|  | 30 | 2 | 65 | 75 | 5 | 307,737 | 501,663,730 | 380,702 | 4,471,702 | 1* | -12 | -2 | -4 | 45682 | - |
|  | 30 | 2 | 45 | 55 | 5 | 298,611 | 1,116,709,025 | 347,950 | 6,172,804 | 0* | -15 | 118 | 33 | 25527 | - |
|  | 30 | 2 | 70 | 80 | 5 | 212,419 | 636,935,832 | 282,176 | 3,042,105 | 1* | -40 | 24 | -34 | 40954 | - |
|  | 30 | 3 | 45 | 81 | 12 | 571,312 | 1,582,020,522 | 690,241 | 6,823,550 | 0* | 63 | 209 | 47 | 66678 | - |
|  | 30 | 3 | 45 | 75 | 10 | 549,025 | 1,403,093,384 | 656,199 | 6,482,618 | 0* | 56 | 174 | 40 | 60176 | - |
|  | 30 | 3 | 50 | 80 | 10 | 536,965 | 1,192,567,897 | 650,613 | 5,978,095 | 0* | 53 | 133 | 29 | 64574 | - |
|  | 30 | 3 | 50 | 74 | 8 | 508,913 | 1,007,716,600 | 609,363 | 5,595,304 | 0* | 45 | 97 | 21 | 57058 | - |
|  | 30 | 3 | 50 | 71 | 7 | 482,100 | 924,248,271 | 573,994 | 5,308,497 | 0* | 37 | 81 | 14 | 51784 | - |
|  | 30 | 3 | 45 | 69 | 8 | 494,506 | 1,242,906,644 | 584,924 | 5,920,596 | 0* | 41 | 143 | 28 | 49881 | - |
|  | 30 | 3 | 55 | 79 | 8 | 474,764 | 884,232,458 | 578,626 | 5,054,431 | 0* | 35 | 73 | 9 | 60163 | - |
|  | 30 | 3 | 55 | 76 | 7 | 461,929 | 785,594,381 | 559,063 | 4,856,016 | 0* | 31 | 53 | 5 | 56423 | - |
|  | 30 | 3 | 45 | 66 | 7 | 453,534 | 1,173,939,824 | 534,339 | 5,525,443 | 0* | 29 | 129 | 19 | 43830 | - |
|  | 30 | 3 | 55 | 70 | 5 | 408,468 | 609,800,187 | 488,270 | 4,231,905 | 1* | 16 | 19 | -9 | 45708 | - |
|  | 30 | 3 | 50 | 65 | 5 | 399,519 | 779,488,505 | 471,727 | 4,481,907 | 1* | 14 | 52 | -3 | 39253 | - |
|  | 30 | 3 | 60 | 81 | 7 | 390,015 | 764,980,767 | 486,404 | 3,949,507 | 1* | 11 | 49 | -15 | 55838 | - |
|  | 30 | 3 | 60 | 75 | 5 | 364,506 | 566,305,643 | 447,694 | 3,550,691 | 1* | 4 | 11 | -23 | 48410 | - |
|  | 30 | 3 | 45 | 60 | 5 | 345,499 | 1,025,467,262 | 405,579 | 4,477,079 | 1* | -2 | 100 | -3 | 30836 | - |
| Policy Position | Cut off (µg Hb/g) | Interval (Years) | Start Age | Stop Age | Number of Lifetime Screens | ΔQALY | ΔC (€) | ΔLYG | Total Number of Colonoscopies | Within Capacity 0= No 1= Yes | % ΔQALY | % ΔC | %ΔNo. Colonoscopies |  |  |
|  | 30 | 3 | 55 | 64 | 3 | 312,866 | 464,666,975 | 371,498 | 3,242,014 | 1* | -11 | -9 | -30 | 31960 | - |
|  | 30 | 3 | 60 | 69 | 3 | 298,580 | 390,857,912 | 362,529 | 2,839,486 | 1* | -15 | -24 | -39 | 36044 | - |
|  | 30 | 3 | 65 | 80 | 5 | 281,256 | 642,805,614 | 363,337 | 2,821,533 | 1* | -20 | 26 | -39 | 46139 | - |
|  | 30 | 3 | 50 | 59 | 3 | 279,287 | 623,678,992 | 329,495 | 3,294,458 | 1* | -21 | 22 | -29 | 25361 | - |
|  | 30 | 3 | 65 | 74 | 3 | 246,693 | 433,903,412 | 313,510 | 2,335,910 | 1* | -30 | -15 | -50 | 37014 | - |
|  | 30 | 3 | 45 | 54 | 3 | 217,246 | 820,101,090 | 256,358 | 3,121,184 | 1* | -38 | 60 | -33 | 18203 | - |
|  | 30 | 3 | 70 | 79 | 3 | 173,815 | 559,350,588 | 238,515 | 1,874,455 | 1* | -51 | 9 | -60 | 34178 | - |
|  | 30 | 4 | 45 | 81 | 9 | 485,172 | 1,381,260,612 | 597,915 | 4,469,030 | 1* | 38 | 170 | -4 | 58339 | - |
|  | 30 | 4 | 45 | 77 | 8 | 476,253 | 1,262,890,869 | 581,654 | 4,296,090 | 1* | 36 | 147 | -7 | 54885 | - |
|  | 30 | 4 | 50 | 82 | 8 | 459,281 | 1,120,375,479 | 569,345 | 4,036,691 | 1* | 31 | 119 | -13 | 57858 | - |
|  | 30 | 4 | 50 | 74 | 6 | 433,635 | 884,373,588 | 527,612 | 3,649,427 | 1* | 23 | 73 | -21 | 49815 | - |
|  | 30 | 4 | 45 | 69 | 6 | 420,441 | 1,049,787,901 | 504,542 | 3,805,979 | 1* | 20 | 105 | -18 | 43107 | - |
|  | 30 | 4 | 55 | 79 | 6 | 407,106 | 812,411,530 | 505,463 | 3,359,131 | 1* | 16 | 59 | -28 | 53008 | - |
|  | 30 | 4 | 50 | 70 | 5 | 401,079 | 776,649,891 | 483,317 | 3,368,980 | 1* | 14 | 52 | -27 | 43271 | - |
|  | 30 | 4 | 55 | 75 | 5 | 391,233 | 689,053,266 | 480,582 | 3,138,590 | 1* | 11 | 35 | -32 | 48273 | - |
|  | 30 | 4 | 55 | 71 | 4 | 360,178 | 576,293,462 | 437,959 | 2,844,056 | 1* | 3 | 13 | -39 | 41676 | - |
|  | 30 | 4 | 45 | 65 | 5 | 372,120 | 964,125,641 | 443,359 | 3,455,207 | 1* | 6 | 88 | -25 | 35783 | - |
|  | 30 | 4 | 50 | 66 | 4 | 354,391 | 685,742,490 | 423,691 | 3,010,991 | 1* | 1 | 34 | -35 | 35802 | - |
|  | 30 | 4 | 60 | 80 | 5 | 348,082 | 742,918,735 | 440,888 | 2,815,030 | 1* | -1 | 45 | -39 | 51481 | - |
|  | 30 | 4 | 60 | 76 | 4 | 320,248 | 558,927,187 | 401,147 | 2,380,326 | 1* | -9 | 9 | -49 | 43954 | - |
|  | 30 | 4 | 55 | 67 | 3 | 315,330 | 480,123,014 | 379,898 | 2,468,629 | 1* | -10 | -6 | -47 | 34088 | - |
|  | 30 | 4 | 60 | 72 | 3 | 291,599 | 439,425,170 | 361,112 | 2,108,221 | 1* | -17 | -14 | -55 | 37473 | - |
|  | 30 | 4 | 45 | 61 | 4 | 309,549 | 881,119,968 | 367,470 | 3,018,571 | 1* | -12 | 72 | -35 | 28060 | - |
|  | 30 | 4 | 50 | 62 | 3 | 292,836 | 602,755,704 | 348,512 | 2,563,145 | 1* | -17 | 18 | -45 | 27796 | - |
|  | 30 | 4 | 65 | 81 | 4 | 245,065 | 644,200,728 | 324,508 | 1,961,845 | 1* | -30 | 26 | -58 | 41672 | - |
|  | 30 | 4 | 65 | 77 | 3 | 234,121 | 510,082,611 | 305,167 | 1,749,797 | 1* | -33 | 0 | -62 | 37576 | - |
|  | 30 | 4 | 45 | 57 | 3 | 239,304 | 782,081,513 | 283,727 | 2,498,539 | 1* | -32 | 53 | -46 | 20563 | - |
|  | 30 | 4 | 70 | 82 | 3 | 158,678 | 627,083,981 | 225,186 | 1,459,539 | 1* | -55 | 22 | -69 | 33200 | - |
|  | 30 | 5 | 45 | 80 | 7 | 423,989 | 1,218,571,847 | 527,834 | 3,216,893 | 1* | 21 | 138 | -31 | 51647 | - |
|  | 30 | 5 | 45 | 75 | 6 | 410,155 | 1,076,287,743 | 504,159 | 3,026,385 | 1* | 17 | 110 | -35 | 46940 | - |
|  | 30 | 5 | 50 | 80 | 6 | 400,903 | 973,934,503 | 500,920 | 2,816,100 | 1* | 14 | 90 | -39 | 50571 | - |
| Policy Position | Cut off (µg Hb/g) | Interval (Years) | Start Age | Stop Age | Number of Lifetime Screens | ΔQALY | ΔC (€) | ΔLYG | Total Number of Colonoscopies | Within Capacity 0= No 1= Yes | % ΔQALY | % ΔC | %ΔNo. Colonoscopies |  |  |
|  | 30 | 5 | 50 | 75 | 5 | 386,707 | 829,028,988 | 476,730 | 2,618,974 | 1* | 10 | 62 | -44 | 45757 | - |
|  | 30 | 5 | 45 | 70 | 5 | 377,314 | 948,600,075 | 458,107 | 2,784,706 | 1* | 7 | 85 | -40 | 39857 | - |
|  | 30 | 5 | 55 | 80 | 5 | 359,767 | 785,705,918 | 453,168 | 2,355,481 | 1* | 2 | 53 | -49 | 48082 | - |
|  | 30 | 5 | 50 | 70 | 4 | 352,598 | 699,827,827 | 428,996 | 2,368,127 | 1* | 0 | 37 | -49 | 38448 | - |
|  | 30 | 5 | 55 | 75 | 4 | 345,140 | 636,414,920 | 428,326 | 2,150,664 | 1* | -2 | 24 | -54 | 43140 | - |
|  | 30 | 5 | 55 | 70 | 3 | 309,785 | 505,081,924 | 378,952 | 1,890,437 | 1* | -12 | -1 | -59 | 35529 | - |
|  | 30 | 5 | 45 | 65 | 4 | 326,293 | 844,769,628 | 391,877 | 2,479,922 | 1* | -7 | 65 | -47 | 31648 | - |
|  | 30 | 5 | 50 | 65 | 3 | 300,208 | 594,348,110 | 361,032 | 2,051,402 | 1* | -15 | 16 | -56 | 29983 | - |
|  | 30 | 5 | 60 | 80 | 4 | 294,955 | 659,360,629 | 379,856 | 1,884,874 | 1* | -16 | 29 | -59 | 43576 | - |
|  | 30 | 5 | 60 | 75 | 3 | 279,756 | 505,329,246 | 353,944 | 1,669,199 | 1* | -20 | -1 | -64 | 38403 | - |
|  | 30 | 5 | 55 | 65 | 2 | 255,084 | 397,186,354 | 308,124 | 1,552,132 | 1* | -27 | -22 | -67 | 26703 | - |
|  | 30 | 5 | 60 | 70 | 2 | 242,720 | 366,222,290 | 302,057 | 1,388,352 | 1* | -31 | -28 | -70 | 30390 | - |
|  | 30 | 5 | 45 | 60 | 3 | 258,297 | 751,697,612 | 308,291 | 2,097,984 | 1* | -26 | 47 | -55 | 23149 | - |
|  | 30 | 5 | 50 | 60 | 2 | 228,359 | 500,061,701 | 272,793 | 1,652,114 | 1* | -35 | -2 | -64 | 21052 | - |
|  | 30 | 5 | 65 | 80 | 3 | 217,507 | 580,727,876 | 290,659 | 1,426,017 | 1* | -38 | 13 | -69 | 37050 | - |
|  | 30 | 5 | 65 | 75 | 2 | 201,048 | 419,752,243 | 262,905 | 1,195,601 | 1* | -43 | -18 | -74 | 31544 | - |
|  | 30 | 5 | 45 | 55 | 2 | 180,286 | 639,903,271 | 215,046 | 1,622,337 | 1* | -49 | 25 | -65 | 15087 | - |
|  | 30 | 5 | 70 | 80 | 2 | 139,987 | 532,381,713 | 199,106 | 1,012,628 | 1* | -60 | 4 | -78 | 28792 | - |
|  | 40 | 1 | 45 | 80 | 35 | 832,619 | 3,052,464,257 | 960,244 | 28,435,120 | 0 | 137 | 496 | 513 | 88954 | - |
|  | 40 | 1 | 50 | 80 | 30 | 789,126 | 2,216,660,058 | 909,514 | 25,324,635 | 0 | 125 | 333 | 446 | 87599 | - |
|  | 40 | 1 | 45 | 75 | 30 | 807,232 | 2,874,636,200 | 928,022 | 27,814,462 | 0 | 130 | 461 | 500 | 83132 | - |
|  | 40 | 1 | 50 | 75 | 25 | 762,211 | 2,025,409,463 | 875,211 | 24,654,523 | 0 | 117 | 296 | 432 | 81370 | - |
|  | 40 | 1 | 55 | 80 | 25 | 705,222 | 1,572,008,525 | 815,670 | 21,201,956 | 0 | 101 | 207 | 357 | 83493 | - |
|  | 40 | 1 | 45 | 70 | 25 | 755,446 | 2,680,188,126 | 866,548 | 26,356,454 | 0 | 115 | 423 | 468 | 74261 | - |
|  | 40 | 1 | 50 | 70 | 20 | 706,080 | 1,818,003,589 | 808,691 | 23,064,263 | 0 | 101 | 255 | 397 | 71760 | - |
|  | 40 | 1 | 55 | 75 | 20 | 675,961 | 1,367,749,639 | 778,528 | 20,470,100 | 0 | 92 | 167 | 341 | 76708 | - |
|  | 40 | 1 | 55 | 70 | 15 | 614,202 | 1,144,177,410 | 705,391 | 18,693,886 | 0 | 75 | 123 | 303 | 66189 | - |
|  | 40 | 1 | 45 | 65 | 20 | 668,489 | 2,461,316,708 | 766,939 | 23,816,360 | 0 | 90 | 381 | 414 | 62553 | - |
|  | 40 | 1 | 50 | 65 | 15 | 612,124 | 1,583,786,993 | 700,862 | 20,254,665 | 0 | 74 | 209 | 337 | 59063 | - |
|  | 40 | 1 | 60 | 80 | 20 | 570,830 | 1,155,392,724 | 670,782 | 16,418,853 | 0 | 62 | 126 | 254 | 75007 | - |
|  | 40 | 1 | 60 | 75 | 15 | 538,114 | 934,065,637 | 629,371 | 15,594,848 | 0 | 53 | 82 | 236 | 67477 | - |
| Policy Position | Cut off (µg Hb/g) | Interval (Years) | Start Age | Stop Age | Number of Lifetime Screens | ΔQALY | ΔC (€) | ΔLYG | Total Number of Colonoscopies | Within Capacity 0= No 1= Yes | % ΔQALY | % ΔC | %ΔNo. Colonoscopies |  |  |
|  | 40 | 1 | 55 | 65 | 10 | 507,610 | 908,056,444 | 584,358 | 15,436,071 | 0 | 44 | 77 | 233 | 51834 | - |
|  | 40 | 1 | 45 | 60 | 15 | 543,137 | 2,181,299,070 | 626,233 | 20,025,718 | 0 | 55 | 326 | 332 | 48674 | - |
|  | 40 | 1 | 60 | 70 | 10 | 468,577 | 700,087,152 | 547,562 | 13,505,046 | 0 | 33 | 37 | 191 | 55595 | - |
|  | 40 | 1 | 50 | 60 | 10 | 472,981 | 1,289,888,754 | 545,047 | 15,949,416 | 0 | 35 | 152 | 244 | 43732 | - |
|  | 40 | 1 | 65 | 80 | 15 | 414,622 | 907,530,530 | 501,976 | 11,392,054 | 0 | 18 | 77 | 146 | 63027 | - |
|  | 40 | 1 | 70 | 80 | 10 | 258,567 | 784,410,431 | 332,341 | 6,591,084 | 0* | -26 | 53 | 42 | 48098 | - |
|  | 40 | 2 | 45 | 81 | 18 | 655,958 | 1,987,993,976 | 791,885 | 10,692,841 | 0 | 87 | 288 | 131 | 75522 | - |
|  | 40 | 2 | 50 | 80 | 15 | 616,114 | 1,476,017,951 | 744,324 | 9,272,862 | 0 | 75 | 188 | 100 | 72889 | - |
|  | 40 | 2 | 45 | 75 | 15 | 629,462 | 1,789,408,620 | 752,968 | 10,275,675 | 0 | 79 | 249 | 122 | 68279 | - |
|  | 40 | 2 | 50 | 76 | 13 | 597,929 | 1,337,974,121 | 717,519 | 8,995,656 | 0 | 70 | 161 | 94 | 67868 | - |
|  | 40 | 2 | 45 | 71 | 13 | 592,013 | 1,664,467,966 | 704,295 | 9,761,274 | 0 | 69 | 225 | 110 | 61069 | - |
|  | 40 | 2 | 55 | 81 | 13 | 546,962 | 1,142,842,270 | 667,229 | 7,823,192 | 0 | 56 | 123 | 69 | 69728 | - |
|  | 40 | 2 | 50 | 70 | 10 | 539,402 | 1,138,526,000 | 641,262 | 8,196,846 | 0 | 54 | 122 | 77 | 56609 | - |
| Planned age expansion (4) | 40 | 2 | 55 | 75 | 10 | 517,847 | 931,519,640 | 624,837 | 7,353,689 | 0 | 47 | 82 | 59 | 61825 | - |
|  | 40 | 2 | 55 | 71 | 8 | 476,397 | 794,499,796 | 571,042 | 6,765,404 | 0* | 36 | 55 | 46 | 53821 | - |
|  | 40 | 2 | 45 | 65 | 10 | 501,981 | 1,482,074,552 | 593,623 | 8,540,432 | 0 | 43 | 189 | 84 | 47830 | - |
|  | 40 | 2 | 50 | 66 | 8 | 477,087 | 1,016,536,462 | 564,701 | 7,336,756 | 0* | 36 | 99 | 58 | 47380 | - |
|  | 40 | 2 | 60 | 80 | 10 | 440,117 | 870,995,011 | 546,537 | 5,921,862 | 0* | 25 | 70 | 28 | 61790 | - |
|  | 40 | 2 | 60 | 76 | 8 | 419,271 | 722,327,245 | 516,347 | 5,603,145 | 0* | 19 | 41 | 21 | 56114 | - |
|  | 40 | 2 | 45 | 61 | 8 | 419,106 | 1,349,638,689 | 495,535 | 7,393,963 | 0 | 19 | 164 | 59 | 38075 | - |
|  | 40 | 2 | 55 | 65 | 5 | 373,877 | 607,865,205 | 445,859 | 5,321,750 | 0* | 6 | 19 | 15 | 38812 | - |
| Current Policy (3) | 40 | 2 | 60 | 70 | 5 | 351,316 | 512,011,709 | 428,784 | 4,637,215 | 0* | 0 | 0 | 0 | 43142 | - |
|  | 40 | 2 | 50 | 60 | 5 | 343,514 | 814,710,211 | 406,885 | 5,486,305 | 0* | -2 | 59 | 18 | 31679 | - |
|  | 40 | 2 | 65 | 81 | 8 | 321,310 | 779,250,054 | 414,845 | 4,319,051 | 1* | -9 | 52 | -7 | 52830 | - |
|  | 40 | 2 | 65 | 75 | 5 | 286,568 | 546,414,128 | 365,124 | 3,740,782 | 1* | -18 | 7 | -19 | 43516 | - |
|  | 40 | 2 | 45 | 55 | 5 | 272,661 | 1,079,412,159 | 324,500 | 5,219,934 | 0* | -22 | 111 | 13 | 23399 | - |
|  | 40 | 2 | 70 | 80 | 5 | 197,579 | 674,327,031 | 271,767 | 2,519,882 | 1* | -44 | 32 | -46 | 39255 | - |
|  | 40 | 3 | 45 | 81 | 12 | 539,089 | 1,602,510,285 | 669,137 | 5,729,585 | 0* | 53 | 213 | 24 | 64622 | - |
|  | 40 | 3 | 45 | 75 | 10 | 516,898 | 1,410,058,606 | 633,712 | 5,429,543 | 0* | 47 | 175 | 17 | 57894 | - |
|  | 40 | 3 | 50 | 80 | 10 | 505,343 | 1,229,592,596 | 628,931 | 4,983,826 | 0* | 44 | 140 | 7 | 62327 | - |
| Policy Position | Cut off (µg Hb/g) | Interval (Years) | Start Age | Stop Age | Number of Lifetime Screens | ΔQALY | ΔC (€) | ΔLYG | Total Number of Colonoscopies | Within Capacity 0= No 1= Yes | % ΔQALY | % ΔC | %ΔNo. Colonoscopies |  |  |
|  | 40 | 3 | 50 | 74 | 8 | 477,375 | 1,032,526,491 | 586,183 | 4,653,247 | 0* | 36 | 102 | 0 | 54582 | - |
|  | 40 | 3 | 50 | 71 | 7 | 451,192 | 941,574,874 | 550,401 | 4,408,724 | 1* | 28 | 84 | -5 | 49282 | - |
|  | 40 | 3 | 45 | 69 | 8 | 463,016 | 1,235,446,436 | 560,816 | 4,945,576 | 0* | 32 | 141 | 7 | 47444 | - |
|  | 40 | 3 | 55 | 79 | 8 | 443,903 | 936,905,948 | 556,889 | 4,198,496 | 1* | 26 | 83 | -9 | 57815 | - |
|  | 40 | 3 | 55 | 76 | 7 | 431,280 | 832,978,007 | 536,853 | 4,025,272 | 1* | 23 | 63 | -13 | 53997 | - |
|  | 40 | 3 | 45 | 66 | 7 | 423,125 | 1,159,224,635 | 510,110 | 4,609,317 | 1* | 20 | 126 | -1 | 41430 | - |
|  | 40 | 3 | 55 | 70 | 5 | 379,329 | 644,186,148 | 465,584 | 3,490,004 | 1* | 8 | 26 | -25 | 43268 | - |
|  | 40 | 3 | 50 | 65 | 5 | 371,057 | 782,842,969 | 448,114 | 3,709,809 | 1* | 6 | 53 | -20 | 36863 | - |
|  | 40 | 3 | 60 | 81 | 7 | 363,893 | 821,934,923 | 467,984 | 3,249,392 | 1* | 4 | 61 | -30 | 53621 | - |
|  | 40 | 3 | 60 | 75 | 5 | 339,074 | 612,208,462 | 428,583 | 2,904,858 | 1* | -3 | 20 | -37 | 46099 | - |
|  | 40 | 3 | 45 | 60 | 5 | 318,675 | 999,157,139 | 382,388 | 3,717,818 | 1* | -9 | 95 | -20 | 28665 | - |
|  | 40 | 3 | 55 | 64 | 3 | 287,464 | 484,951,501 | 350,068 | 2,655,900 | 1* | -18 | -5 | -43 | 29787 | - |
|  | 40 | 3 | 60 | 69 | 3 | 275,394 | 423,943,758 | 343,569 | 2,305,419 | 1* | -22 | -17 | -50 | 33857 | - |
|  | 40 | 3 | 65 | 80 | 5 | 261,622 | 688,879,418 | 349,142 | 2,294,817 | 1* | -26 | 35 | -51 | 44194 | - |
|  | 40 | 3 | 50 | 59 | 3 | 256,930 | 615,085,390 | 309,518 | 2,712,412 | 1* | -27 | 20 | -42 | 23456 | - |
|  | 40 | 3 | 65 | 74 | 3 | 228,254 | 468,104,170 | 299,000 | 1,882,127 | 1* | -35 | -9 | -59 | 35081 | - |
|  | 40 | 3 | 45 | 54 | 3 | 197,941 | 787,461,947 | 238,524 | 2,578,134 | 1* | -44 | 54 | -44 | 16635 | - |
|  | 40 | 3 | 70 | 79 | 3 | 160,501 | 590,579,766 | 228,384 | 1,517,950 | 1* | -54 | 15 | -67 | 32544 | - |
|  | 40 | 4 | 45 | 81 | 9 | 454,871 | 1,399,822,326 | 576,431 | 3,666,678 | 1* | 29 | 173 | -21 | 56230 | - |
|  | 40 | 4 | 45 | 77 | 8 | 446,142 | 1,274,045,233 | 559,629 | 3,515,991 | 1* | 27 | 149 | -24 | 52677 | - |
|  | 40 | 4 | 50 | 82 | 8 | 429,797 | 1,156,866,635 | 548,134 | 3,302,982 | 1* | 22 | 126 | -29 | 55628 | - |
|  | 40 | 4 | 50 | 74 | 6 | 404,790 | 906,502,122 | 505,491 | 2,968,012 | 1* | 15 | 77 | -36 | 47454 | - |
|  | 40 | 4 | 45 | 69 | 6 | 391,619 | 1,044,827,528 | 481,638 | 3,106,093 | 1* | 11 | 104 | -33 | 40834 | - |
|  | 40 | 4 | 55 | 79 | 6 | 378,738 | 857,241,491 | 484,301 | 2,724,363 | 1* | 8 | 67 | -41 | 50658 | - |
|  | 40 | 4 | 50 | 70 | 5 | 373,297 | 790,728,136 | 461,122 | 2,735,129 | 1* | 6 | 54 | -41 | 40946 | - |
|  | 40 | 4 | 55 | 75 | 5 | 363,497 | 726,111,151 | 459,209 | 2,536,331 | 1* | 3 | 42 | -45 | 45918 | - |
|  | 40 | 4 | 55 | 71 | 4 | 333,297 | 606,427,033 | 416,418 | 2,290,112 | 1* | -5 | 18 | -51 | 39331 | - |
|  | 40 | 4 | 45 | 65 | 5 | 345,315 | 950,079,607 | 421,098 | 2,817,972 | 1* | -2 | 86 | -39 | 33642 | - |
|  | 40 | 4 | 50 | 66 | 4 | 328,628 | 691,451,211 | 402,261 | 2,442,137 | 1* | -6 | 35 | -47 | 33626 | - |
|  | 40 | 4 | 60 | 80 | 5 | 324,078 | 792,361,179 | 423,436 | 2,271,907 | 1* | -8 | 55 | -51 | 49378 | - |
|  | 40 | 4 | 60 | 76 | 4 | 296,952 | 598,975,325 | 383,099 | 1,896,691 | 1* | -15 | 17 | -59 | 41814 | - |
| Policy Position | Cut off (µg Hb/g) | Interval (Years) | Start Age | Stop Age | Number of Lifetime Screens | ΔQALY | ΔC (€) | ΔLYG | Total Number of Colonoscopies | Within Capacity<br>0= No<br>1= Yes | % ΔQALY | % ΔC | %ΔNo. Colonoscopies |  |  |
|  | 40 | 4 | 55 | 67 | 3 | 290,608 | 502,059,880 | 359,287 | 1,981,724 | 1* | -17 | -2 | -57 | 31916 | - |
|  | 40 | 4 | 60 | 72 | 3 | 269,132 | 473,091,308 | 342,921 | 1,672,010 | 1* | -23 | -8 | -64 | 35353 | - |
|  | 40 | 4 | 45 | 61 | 4 | 285,804 | 858,098,279 | 346,857 | 2,461,120 | 1* | -19 | 68 | -47 | 26182 | - |
|  | 40 | 4 | 50 | 62 | 3 | 269,963 | 600,842,580 | 328,659 | 2,076,177 | 1* | -23 | 17 | -55 | 25873 | - |
|  | 40 | 4 | 65 | 81 | 4 | 227,009 | 684,666,986 | 310,930 | 1,566,126 | 1* | -35 | 34 | -66 | 39810 | - |
|  | 40 | 4 | 65 | 77 | 3 | 216,524 | 545,092,940 | 291,432 | 1,384,721 | 1* | -38 | 6 | -70 | 35709 | - |
|  | 40 | 4 | 45 | 57 | 3 | 219,256 | 753,879,981 | 265,576 | 2,035,451 | 1* | -38 | 47 | -56 | 18961 | - |
|  | 40 | 4 | 70 | 82 | 3 | 146,274 | 656,767,988 | 215,686 | 1,167,618 | 1* | -58 | 28 | -75 | 31655 | - |
|  | 40 | 5 | 45 | 80 | 7 | 395,518 | 1,230,204,791 | 506,268 | 2,585,504 | 1* | 13 | 140 | -44 | 49495 | - |
|  | 40 | 5 | 45 | 75 | 6 | 382,070 | 1,080,653,063 | 482,210 | 2,423,387 | 1* | 9 | 111 | -48 | 44749 | - |
|  | 40 | 5 | 50 | 80 | 6 | 373,785 | 999,592,131 | 480,185 | 2,246,826 | 1* | 6 | 95 | -52 | 48377 | - |
|  | 40 | 5 | 50 | 75 | 5 | 360,307 | 847,525,345 | 455,966 | 2,080,270 | 1* | 3 | 66 | -55 | 43579 | - |
|  | 40 | 5 | 45 | 70 | 5 | 350,153 | 944,507,885 | 435,932 | 2,226,717 | 1* | 0 | 84 | -52 | 37694 | - |
|  | 40 | 5 | 55 | 80 | 5 | 333,703 | 824,410,604 | 433,204 | 1,864,114 | 1* | -5 | 61 | -60 | 45884 | - |
|  | 40 | 5 | 50 | 70 | 4 | 326,964 | 710,812,937 | 407,923 | 1,876,218 | 1* | -7 | 39 | -60 | 36286 | - |
|  | 40 | 5 | 55 | 75 | 4 | 319,785 | 668,397,221 | 408,296 | 1,691,016 | 1* | -9 | 31 | -64 | 40948 | - |
|  | 40 | 5 | 45 | 65 | 4 | 301,780 | 830,958,436 | 371,026 | 1,986,417 | 1* | -14 | 62 | -57 | 29684 | - |
|  | 40 | 5 | 55 | 70 | 3 | 285,855 | 528,857,979 | 359,263 | 1,481,388 | 1* | -19 | 3 | -68 | 33443 | - |
|  | 40 | 5 | 50 | 65 | 3 | 277,105 | 596,751,993 | 341,253 | 1,625,619 | 1* | -21 | 17 | -65 | 28041 | - |
|  | 40 | 5 | 60 | 80 | 4 | 273,236 | 698,814,185 | 362,956 | 1,480,334 | 1* | -22 | 36 | -68 | 41518 | - |
|  | 40 | 5 | 60 | 75 | 3 | 258,702 | 539,048,464 | 337,011 | 1,298,675 | 1* | -26 | 5 | -72 | 36378 | - |
|  | 40 | 5 | 55 | 65 | 2 | 234,236 | 410,994,541 | 290,203 | 1,214,760 | 1* | -33 | -20 | -74 | 24897 | - |
|  | 40 | 5 | 60 | 70 | 2 | 223,496 | 392,251,527 | 285,946 | 1,073,511 | 1* | -36 | -23 | -77 | 28537 | - |
|  | 40 | 5 | 45 | 60 | 3 | 237,425 | 729,472,599 | 289,707 | 1,683,848 | 1* | -32 | 42 | -64 | 21477 | - |
|  | 40 | 5 | 50 | 60 | 2 | 209,762 | 493,166,843 | 256,132 | 1,312,452 | 1* | -40 | -4 | -72 | 19507 | - |
|  | 40 | 5 | 65 | 80 | 3 | 200,769 | 613,901,647 | 277,464 | 1,112,573 | 1* | -43 | 20 | -76 | 35246 | - |
|  | 40 | 5 | 65 | 75 | 2 | 185,289 | 447,436,913 | 250,036 | 920,320 | 1* | -47 | -13 | -80 | 29815 | - |
|  | 40 | 5 | 45 | 55 | 2 | 164,407 | 611,938,881 | 200,307 | 1,304,021 | 1* | -53 | 20 | -72 | 13838 | - |
|  | 40 | 5 | 70 | 80 | 2 | 128,496 | 556,541,437 | 189,832 | 789,794 | 1* | -63 | 9 | -83 | 27297 | - |

\*= within the future extended capacity

